# Oral Blarcamesine Phase IIb/III Trial Confirms Identified Precision Medicine Patient Population – Significant Broad Clinical and Quality of Life Improvements for Early Alzheimer’s Disease Patients

**DOI:** 10.1101/2025.09.27.25336656

**Authors:** Stephen Macfarlane, Timo Grimmer, Ken Teo, Terence J O’Brien, Michael Woodward, Jennifer Grunfeld, Alastair Mander, Bruce J. Brew, Philip Morris, Cathy Short, Susan Kurrle, Rosalyn Lai, Sneha Bharadwaj, Peter Drysdale, Jonathan Sturm, Simon J.G. Lewis, Chris Kalafatis, Saif Sharif, Nicholas Mannering, J. Emer MacSweeney, Stephen Pearson, Craig Evans, Neel Bhatt, Sandra Connell, Jennifer Lynch, Paul LJ Dautzenberg, Niels Prins, Lutz Frölich, Pawel Tacik, Oliver Peters, Alexandre Henri-Bhargava, Stephen H Pasternak, Andrew Frank, Howard Chertkow, Jennifer Ingram, Ging-Yuek Robin Hsiung, Carmela Tartaglia, Sharon Cohen, Olivier Courrèges, Luca M Villa, Elizabeth Gordon, Nicolas Guizard, Jeffrey Edwards, Terrie Kellmeyer, Juan Carlos Lopez-Talavera, David S Gould, Wolfgang Liedtke, Kun Jin, William R Chezem, Christopher U Missling, Audrey Gabelle, Marwan N Sabbagh

## Abstract

**IMPORTANCE:** There are no approved oral disease-modifying treatments for Alzheimer’s disease (AD) with the ability to prolong time in a stable disease state and with clinically meaningful outcomes clear to patients and caregivers.

**DESIGN:** The Phase IIb/III ANAVEX2-73-AD-004 study was a randomized, double-blind, placebo-controlled, 48-week trial with prespecified gene or GWAS identified genetic variant populations related to the mechanism of the pharmacological intervention.

**OBJECTIVE:** Providing evidence of improved Precision Medicine neurology clinical treatment responses with optimal blarcamesine dose for up to ∼70% of AD patients within prespecified SIGMAR1 (ABCLEAR1) and GWAS identified COL24A1 (ABCLEAR2) and SIGMAR1/COL24A1 (ABCLEAR3) non-missense gene populations. Describing once-daily, oral therapeutic intervention of blarcamesine in early AD, with a differentiated upstream and constitutional mechanism of action by enhancing autophagy through SIGMAR1 activation and restoration of cellular homeostasis.

**SETTING:** Multicenter - 52 medical research centers/hospitals in 5 countries.

**INTERVENTION:** 508 participants with Early AD (Stage 3) were randomized to receive blarcamesine (n = 338) oral capsules once daily either in medium dose group (30 mg) or in high dose group (50 mg) or placebo (n = 170) for 48 weeks. An open-label-extension study ATTENTION-AD, continued for up to 192 weeks.

**MAIN OUTCOME AND MEASURES:** Further improved clinical and biomarker outcomes of the ABCLEAR2 and ABCLEAR3 populations were accretive to the previously reported intent-to-treat (ITT) and prespecified ABCLEAR1 populations. The co-primary cognitive and functional outcomes were assessed as changes in ADAS-Cog13 and ADCS-ADL from baseline to 48 weeks. The outcomes include the secondary outcome CDR-SB as well as patient assessed clinical outcomes CGI-I, NPI-Q and QoL-AD (Quality of Life AD Patient) as well as biomarkers global brain volume changes measured by MRI. All clinical endpoints were analyzed using mixed model for repeated measures (MMRM), and volumetric MRI scans were analyzed by general linear model.

**RESULTS:** The ITT population (mean age, 73.7 years; 225 [48.7%] women), consisted of 462 randomized participants, with ABCLEAR1 population comprising of 300 participants, ABCLEAR2 population of 336 participants and ABCLEAR3 of 222 participants. In both the ABCLEAR2 and ABCLEAR3 populations, the co-primary outcomes as well as all other clinical outcomes were statistically significant. ABCLEAR3 blarcamesine group vs. placebo at Week 48 (ADAS-Cog13 difference of −4.179 [95% CI −6.512, −1.845]; P=0.0005; ADCS-ADL difference of +3.131 [95%CI 0.720, 5.542]; P=0.0111; CDR-SB difference of −1.076 [95% CI −1.645, −0.508]; P=0.0002; QoL-AD Patient improvement from baseline of 0.334 [95% CI −1.164, 1.833] and difference of 1.848 [95% CI 0.455, 3.241]; P=0.0095). The clinical outcomes were strongest in the blarcamesine 30 mg cohort (ADAS-Cog13 difference of −4.739 [95% CI −7.370, −2.108]; P=0.0004; ADCS-ADL difference of +4.245 [95%CI 1.518, 6.972]; P=0.0024; CDR-SB difference of −1.414 [95% CI −2.054, −0.775]; P<0.0001; QoL-AD Patient improvement from baseline of 0.182 [95% CI −1.472, 1.835]; and difference of 1.651 [95% CI 0.455, 3.241]; P=0.0392). Whole brain volume loss in blarcamesine group vs. placebo was significantly further decreased from ITT population (37.6%, P=0.0019) to ABCLEAR3 population (44.5%, P=0.0019). Participants in the 30 mg group ABCLEAR3 full safety population with ≥1 serious treatment-emergent adverse events (TEAEs) occurred in 10 participants (12.7%) in the blarcamesine and 6 (9.1%) in the placebo group. Common TEAEs included dizziness, which was transient and mostly mild to moderate in severity. There were no deaths in the blarcamesine group and 1 in the placebo group.

**CONCLUSIONS AND RELEVANCE:** Blarcamesine group demonstrated a balanced safety profile with no associated neuroimaging adverse events. The respective once-daily oral 30 mg blarcamesine cohort in the respective ABCLEAR1, ABCLEAR2, and ABCLEAR3 populations demonstrates further improvement of the already adequate safety profile of the ITT population and hence representing the dose with the most balanced benefit-to-risk ratio. In both ABCLEAR2 and ABCLEAR3 populations significant slowing in decline and even stabilization of clinical worsening was demonstrated. Blarcamesine group vs. placebo was 75.9% reduction in decline at 48 weeks in the total blarcamesine group and was 84.7% in the 30 mg dose group, respectively on the prespecified co-primary cognitive endpoint ADAS-Cog13 in the ABCLEAR3 population. Blarcamesine demonstrated consistently significantly improved clinical effect for all clinical endpoints, which was in accordance with significant and further reduced brain atrophy. Furthermore, a significant absolute improvement in Quality of Life (QoL-AD) scores indicating a reversal of negative trajectory for Alzheimer’s disease patients from baseline to end of trial was observed. The Phase IIb/III ANAVEX2-73-AD-004 clinical study confirmed blarcamesine’s consistent efficacy leading to improved cognitive stabilization with continued benefit in the open-label-extension study ATTENTION-AD up to 192 weeks. Evidenced by the Precision Medicine paradigm, including in a prespecified ABCLEAR1 population, a consistent significant improvement for all clinical endpoints in the ABCLEAR2 and ABCLEAR3 populations, respectively, was demonstrated. Coupled with a convenient once daily, oral pill administration, blarcamesine could represent a novel treatment option for up to ∼70% of early AD patients benefiting from further improved outcomes using directed Precision Medicine to alleviate significant medical and economic burden.

**TRIAL REGISTRATION:** Clinicaltrials.gov: NCT03790709; Open-label extension NCT04314934

**FUNDING:** This work was funded by Anavex Life Sciences.

**Key Message:** Within a heterogeneous Alzheimer’s disease (AD) population, clinical utility of disease-modifying drug candidates could be enhanced via a Precision Medicine approach of treating those with target-relevant genetic profiles, excluding missense gene populations. This effective targeting could alleviate significant medical and economic burden.

## INTRODUCTION

Alzheimer’s disease (AD) remains one of the most expensive debilitating diseases today with a significant economic burden worldwide [1]. By 2050, 1 in 85 people worldwide will be diagnosed with Alzheimer’s disease (AD)[2]. At current estimates, approximately 60 million persons are living with dementia worldwide [1], and this represents a huge healthcare burden on patients, families and health systems worldwide. AD constitutes an estimated 60-80% of all dementias [3]. In the United States alone, health care and long-term care for people with AD and other dementias are projected to reach $1 trillion by 2050 (in 2023 dollars) [4]. The Global economic burden of AD will likely reach $4 trillion - $5 trillion by 2050 [4]. Despite recent regulatory approvals of monoclonal antibodies targeting amyloid-β, limitations in their efficacy, safety (e.g., ARIA risk), and accessibility (e.g., intravenous administration) leave a significant gap in clinical care. Moreover, these therapies often adopt a one-size-fits-all approach, overlooking the biological and genetic heterogeneity that defines AD pathogenesis and therapeutic response. Emerging insights from genomics and systems biology increasingly point to AD as a polygenic, multifactorial disease with distinct molecular subtypes. This heterogeneity underpins variable treatment responses, mandating a shift toward precision medicine approaches that match therapies to genetically and biologically defined patient populations. Within this context, pharmacogenetic targeting based on receptor biology and GWAS-derived markers represents a transformative strategy for AD drug development. In analogy to improved Precision Medicine oncology treatments, the heterogeneity of AD means that within the AD patient populations there could be participants with specific genetic profiles responding more favorably than others to specific prespecified or with GWAS identified genetic variants [5, 17] related to the mechanism of the pharmacological intervention.

Precision medicine aims to redefine neurodegenerative disease treatment by tailoring interventions to individual genetic, molecular, and phenotypic profiles, acknowledging the biological heterogeneity underlying disease progression and treatment response. Emerging therapeutic frameworks in AD integrate genomic, imaging, and fluid biomarkers to create a multi-dimensional patient profile, enabling a shift from symptomatic treatment to true mechanism-based disease modification. It is increasingly appreciated that further upstream, additional, concurrent pathologies other than aggregated β-amyloid or fibrillary tangle tau appear to propagate AD, like impairment of autophagy and that these mechanisms may be more fundamental or causative. Autophagy is an upstream mechanism which allows for the regular, endogenous clearing of protein aggregates under normal conditions. Impairment of autophagy precedes both amyloid beta and tau tangles, and therefore precedes the neurodegenerative process in AD[6, 7]. Autophagy is mediated by the SIGMAR1 (S1R) receptor, an intracellular membrane protein located in the endoplasmic reticulum which induces autophagy when activated [8], and impairs autophagy when ablated [9].SIGMAR1 receptors are widely expressed in the brain [6, 7], and previously published clinical trial data [10] of SIGMAR1 agonists such as blarcamesine showed a slowing of neurodegeneration [11–14]. Additionally, live *in vivo* real-time imaging studies demonstrate how microglial inflammation, which is involved in devastating neurodegenerative diseases, can be resolved by SIGMAR1 activation. The role of SIGMAR1 activation is to reduce the negative response caused by chronic microglia over-activation, which leads to widespread cell death and multiple neurodegenerative diseases, including AD. Thus, SIGMAR1 activation efficiently “switches off” the damaging chronic microglial reaction. Additionally, the action of SIGMAR1 on microglia was found to be specific to damage responses as the response did not affect the normal microglia activity within the brain [15]. Activation of SIGMAR1 in the central nervous system influences the formation of its oligomeric state, promoting interactions with various proteins to produce biological effects [16–18]. Activation of SIGMAR1 also supports pathways that promote cell survival, including mitochondrial health [19], lipid metabolism [11], and stress responses in the endoplasmic reticulum [12], all of which are relevant to neurodegenerative disease mechanisms. Drug candidates with these constitutional features could represent an important new class of AD treatments.

Blarcamesine (ANAVEX®2-73) is an oral investigative medication aiming to return cellular balance by restoring autophagy through SIGMAR1 activation. Blarcamesine has shown promise in enhancing immune function in elderly individuals by promoting autophagy, a process for cellular waste clearance [8].

There are ample precedents (such as *APOE* ε4) for not only genetically predetermined disease susceptibility but also response to treatment [20]. Below, we describe and contrast clinical efficacy data from the entire ITT blarcamesine study population and the respective two target dose cohorts with that of two Wild Type (WT) populations, excluding the respective missense gene mutations. The first population is the previously described, and in the clinical study prespecified SIGMAR1 WT population (homozygous T/T genotype), which has a global frequency of 70.7% [21], excluding the respective homozygous missense gene mutation, SIGMAR1 (rs1800866). The previously reported study demonstrated that blarcamesine responses were more accentuated in this population, excluding the respective missense gene mutation, indicating a target selectivity of blarcamesine and pharmacogenetic ratification of its clinical effect. In the study protocol preplanned unbiased GWAS analysis identified a second, additionally significant genetic WT marker of positive blarcamesine response, also related to autophagy, the COL24A1 WT population.

The COL24A1 WT population, with a global frequency of 71.7% [21], excluding the respective missense gene mutation COL24A1 (rs60891279), demonstrated enhanced response to blarcamesine treatment allowing unimpaired autophagy restoration through stabilization of the ECM (extracellular matrix) in the brain. COL24A1, is expressed by neurons throughout the human brain [22], with highest expression levels in the hippocampal formation and cortex. COL24A1 WT carriers are more responsive to blarcamesine vs its mutant histidine polymorphism at position 641 (=arginine in WT). [23][24] Further details of the mechanism of COL24A1 will be addressed in a separate publication.

We termed the population with beneficial Wild Type (WT) genes as ABCLEAR1 [26] (excluding the SIGMAR1 missense mutation), ABCLEAR2 (excluding the COL24A1 missense mutation) and ABCLEAR3 (excluding both SIGMAR1 and COL24A1 missense mutations) with ‘ABCLEAR’ standing for ‘Alzheimer’s Blarcamesine Cognition Efficacy and Resilience Genes’.

## METHODS

### Study Design

The ANAVEX2-73-AD-004 trial enrolled 508 participants with early AD, was a Phase IIb/III 48-week randomized, double-blind placebo-controlled, multicenter, international trial of blarcamesine in early AD. After completion of the placebo-controlled 48-week study, the majority of participants enrolled into a 96/144-week open label extension (OLE) study, ATTENTION-AD (ClinicalTrials.gov Identifier NCT04314934), completed in June 2024. The 48-week study was conducted at 52 sites across 5 countries; Australia (19 sites), Canada (10 sites), Germany (5 sites), Netherlands (3 sites) and United Kingdom (15 sites) (ClinicalTrials.gov Identifier: NCT03790709) [10]. An independent data and safety monitoring board reviewed safety data periodically throughout the study. The study was conducted in accordance with the Declaration of Helsinki, the International Conference on Harmonization Good Clinical Practice Guidelines, and local regulatory and ethics requirements.

### Participants

Patients aged 60-85 who met the National Institute on Aging (NIA) – Alzheimer’s Association 2011 criteria for diagnosis of early-stage mild dementia due to AD or mild cognitive impairment due to AD [29–31] were eligible to participate, with ≥1 additional criterion required to support a diagnosis of AD: (a) historic or current record of CSF assessment compatible with AD, cut off values of amyloid beta (Aβ)42 <1054 pg/mL, total Tau (tTau) >213 pg/mL, phosphorylated Tau (pTau) >21.3 pg/mL, and Aβ42/Aβ40 ratio <0.064 or CSF pTau181 >27 pg/mL (irrespective of the Aβ42/Aβ40 ratio) by automated Elecsys® CSF or comparable biomarker assay (Roche Diagnostics), or (b) PET scan (amyloid scan or FDG-PET) within 36 months preceding screening, or (c) CT or MRI in past 18 months consistent with a diagnosis of AD [27–29]. A Mini-Mental State Examination (MMSE) score of 20-28 at the screening and randomization visits [30] and a Free and Cued Selective Reminding Test (FCSRT) recall score of ≤17 or total recall score <40 were also required [31, 32]. Patients on acetylcholinesterase inhibitors or other cognitive enhancing medications were required to remain on stable doses for at least 90 days prior to screening.

Study outcome measures were obtained at baseline study entry and at weeks 12, 24, 36 and 48. MRI assessments and blood draws for pathophysiologic biomarkers were obtained at baseline and Week 48. Randomization and intervention and sample size calculation were consistent with the previously described ITT and ABCLEAR1 populations [10].

### Outcomes

#### Clinical endpoints

The co-primary outcomes were reduction in cognitive decline assessed from baseline over 48 weeks with blarcamesine compared to placebo using the 13-item Alzheimer Disease Assessment Scale-Cognition (ADAS-Cog13), and reduction in decline of the ability to perform daily activities assessed from baseline over 48 weeks with blarcamesine compared to placebo using the Alzheimer’s Disease Cooperative Study – Activities of Daily Living (ADCS-ADL) Scale [10].

A score change >2.0 in ADAS-Cog13 is considered to be the clinically significant threshold [33] and while its use as a single endpoint for early AD may be sufficient for regulatory review [34] the co-primary endpoint of ADCS-ADL was also assessed as it is well recognized [34, 35]. The secondary outcome was the reduction in cognitive and functional decline assessed from baseline over 48 weeks with blarcamesine compared with placebo using the Clinical Dementia Rating Scale Sum of Boxes (CDR-SB) [33, 34, 36].

Specified exploratory clinical endpoints included the Mini-Mental State Examination (MMSE), a standardized measure of cognitive impairment, the questionnaire-based Clinical Global Impression – Improvement (CGI-I) scale, a well validated tool across heterogenous populations [37], the NPI-Q (Neuropsychiatric Inventory Questionnaire), providing a reliable, informant-based assessment of neuropsychiatric symptoms of the participant and associated caregiver distress [38], and the patient-assessed endpoint of QoL-AD, a broad personal measure of AD patients’ quality of life, measuring:

`o` Physical health: Overall physical well-being. Energy: Level of energy and vitality.
`o` Mood: Emotional state and feelings.
`o` Living situation: Satisfaction with where the person lives.
`o` Memory: Cognitive function and memory abilities.
`o` Family: Quality of relationships with family members.
`o` Marriage/Significant other: Satisfaction with the relationship with a partner.
`o` Friends: Quality of social relationships with friends.
`o` Self as a whole: Overall self-perception and self-esteem.
`o` Ability to do chores: Capacity to perform household tasks.
`o` Ability to do things for fun: Enjoyment of leisure activities.
`o` Money: Financial well-being.
`o` Life as a whole: Overall satisfaction with life

#### MRI biomarker endpoints

In oncology, reliable biomarkers of disease progression are typically reflected by tumor growth, whereby malignant cells proliferate uncontrollably. By analogy, a pathological hallmark of Alzheimer’s disease (AD) is progressive cerebral atrophy, in which neuronal loss results in measurable reductions in brain volume. The utility of brain atrophy as a biomarker endpoint was assessed for the two new populations, ABCLEAR2 and ABCLEAR3, and compared with findings previously analyzed and reported in the ITT population [10]. Structural MRI was acquired at both baseline and Week 48 to quantify longitudinal changes in brain volume over the course of the study. Volumetric analyzes were conducted on 3D T1-weighted sequences. Volumes of whole brain, total white matter, total grey matter, and lateral ventricles were quantified and analyzed in terms of annualized percent change from baseline volume. Efficacy was evaluated as the reduction of brain volume loss compared to placebo, while efficacy for lateral ventricles was defined as a reduction in the rate of ventricular expansion compared to placebo.

#### SIGMAR1 Gene Genotyping [Common SIGMAR1 WT gene and missense mutation]

As a prespecified exploratory endpoint of the study, clinical efficacy measurements were compared for populations based on absence or presence of the SIGMAR1 gene missense mutation (rs1800866 T > G variant) to assess the impact of the exclusion of this genetic mutation on clinical efficacy. The vast majority of the general population (70.7%) carries the common SIGMAR1 WT = Wild Type gene, whereas a minority of the general population (∼29%) carries the missense polymorphism [glutamine 0002 to proline (Q2P)] mutated SIGMAR1 (rs1800866) gene. [43] We term this SIGMAR1 WT population the ABCLEAR1 population. The WT (Wild Type) homozygous populations SIGMAR1 can be conveniently established with a swab or blood prick or blood test by using a TaqMan qPCR genotyping test, which is similar to those used for APOE4 genotyping which identify the three different alleles of the APOE gene: ε2, ε3, and ε4.

#### COL24A1 Gene Genotyping [Common COL24A1 WT gene and missense mutation]

Unbiased GWAS analysis identified a second, additionally significant genetic marker of positive blarcamesine response emanating from this study. Detailed methodology and results of this GWAS are described in a companion manuscript; see our GWAS for enhanced response to blarcamesine manuscript for more detailed results and pathobiological context of the COL24A1 gene variant which we discovered. Clinical efficacy measurements were compared for populations based on absence or presence of a COL24A1 gene variant to assess the impact of this genetic variant on clinical efficacy. The vast majority of the general population (71.7%) carries the common COL24A1 WT = Wild Type gene, whereas a minority of the general population (∼28%) carries the missense polymorphism [arginine 641 to histidine (R641H)] mutated COL24A1 (rs60891279) gene. [43] The COL24A1 WT population is termed the ABCLEAR2 population. The combined COL24A1 WT and SIGMAR1 WT population, represents the ABCLEAR3 population. Both Wild Type homozygous populations SIGMAR1 WT and COL24A1 WT can be conveniently established with a simple swab or blood prick or blood test. The cost and the turnaround time for ABCLEAR genotyping (e.g., APOE4 genotyping) can be easily integrated into already existing routine clinical standard procedures or workflows.

### Statistical Analyses

Statistical analyses were done with SAS version 9.4 (SAS Institute) or R Project version 4.2.3 (R Foundation).

#### Analysis of Clinical Endpoints

The study protocol prespecified the reduction in decline assessed from baseline over 48 weeks with blarcamesine compared to placebo for the respective co-primary (ADAS-Cog13 and ADCS-ADL) and secondary (CSR-SB) endpoints using the mixed effects model. Hence, all prespecified clinical endpoints, including ADAS-Cog13, ADCS-ADL, CDR-SB, MMSE, CGI-I, NPI-Q and QoL-AD were analyzed using a linear mixed model (mixed model for repeated measures; MMRM). The MMRM analysis method is the convention used for regulatory filings and was used as the primary analysis method in all recent regulatory decisions for aducanumab [39, 40] and lecanemab [41], as well as donanemab [42] with similar specifications.

Primary and secondary analyses were carried out in the protocol-specified analysis population, the “intent-to-treat” (ITT) population, which corresponds to what is typically termed “modified intent-to-treat” (mITT) and was defined as all randomized patients who received at least one study dose and had at least one post-dose clinical measurement.

The change of clinical scores from baseline to Week 48 and respective analysis of MRI biomarkers were analyzed for the two new populations, ABCLEAR2 and ABCLEAR3, consistent with the previously analyzed ITT and ABCLEAR1 populations [10].

#### Safety Objectives - Adverse Events

Safety objectives were evaluated by the incidence of AEs and serious AEs in the full safety population for both active and placebo groups and were summarized for the respective ABCLEAR2 and ABCLEAR3 populations according to event frequency by treatment assignment.

#### Sensitivity Analysis

The primary analyses assume that missing efficacy assessments are missing at random (MAR). The aim of this sensitivity analysis was to evaluate the sensitivity and robustness of the primary efficacy results to deviations from the missing at random (MAR) assumption, by determining the extent of data bias required under a missing not at random (MNAR) scenario to nullify the statistical significance of the treatment effect. A tipping point analysis under missing not at random (MNAR) assumption was conducted for ADAS-Cog13. In this analysis, 100 datasets were first generated with assumptions of MAR using SAS PROC MI. The missing not at random was realized by worsening imputed values in the active arm with increment of 0.02. or by improving imputed values in placebo arm with increment of 0.04 for the patients imputed under MAR. The primary MMRM model was applied to each of the newly imputed datasets. With each incremental change, these results from imputed data were combined using Rubin’s combination rules, with SAS PROV MIANALYZE. The process stops when the primary model result is no longer significant.

## RESULTS

Of 508 participants enrolled, there were 462 randomized participants in the ITT population (mean age 73.7 years), of which 338 completed the trial. The ITT population was randomized to blarcamesine group [n = 298; 30 mg (n = 154) or 50 mg dose group (n = 144), or placebo (n = 164)]. Most enrolled participants had early AD (Stage 3) with baseline MMSE score 20-28, with majority on background anticholinesterase and/or memantine therapy. Baseline AD status was also evidenced by elevated plasma p-Tau (181) and p-Tau (231) at baseline. The ABCLEAR1 population consisted of 300 participants, ABCLEAR2 population of 336 participants and ABCLEAR3 of 222 participants. In both the ABCLEAR2 and ABCLEAR3 populations, the co-primary outcomes as well as all other clinical outcomes were statistically significant. The ABCLEAR2 and ABCLEAR3 populations were similarly balanced at baseline (Table 1).

**Table 1:** Baseline Demographics – ITT and ABCLEAR Populations.

|  |  | ITT Population |  |  |  | ABCCLEAR1 Population |  |  |  | ABCCLEAR2 Population |  |  |  | ABCCLEAR3 Population |  |  |  |
| --- | --- | --- | --- | --- | --- | --- | --- | --- | --- | --- | --- | --- | --- | --- | --- | --- | --- |
|  |  | Blarcamesine<br>30 mg<br>(N=154) | Blarcamesine<br>50 mg<br>(N=144) | Blarcamesine<br>Total<br>(N=298) | Placebo<br>(N=164) | Blarcamesine<br>30 mg<br>(N=106) | Blarcamesine<br>50 mg<br>(N=93) | Blarcamesine<br>Total<br>(N=199) | Placebo<br>(N=101) | Blarcamesine<br>30 mg<br>(N=111) | Blarcamesine<br>50 mg<br>(N=117) | Blarcamesine<br>Total<br>(N=228) | Placebo<br>(N=108) | Blarcamesine<br>30 mg<br>(N=79) | Blarcamesine<br>50 mg<br>(N=77) | Blarcamesine<br>Total<br>(N=156) | Placebo<br>(N=66) |
| Sex, n (%) |  |  |  |  |  |  |  |  |  |  |  |  |  |  |  |  |  |
| Female |  | 74 (48.1) | 69 (47.9) | 143 (48.0) | 82 (50.0) | 53 (50.0) | 45 (48.4) | 98 (49.2) | 52 (51.5) | 52 (46.8) | 62 (53.0) | 114 (50.0) | 50 (46.3) | 40 (50.6) | 41 (53.2) | 81 (51.9) | 33 (50.0) |
| Male |  | 80 (51.9) | 75 (52.1) | 155 (52.0) | 82 (50.0) | 53 (50.0) | 48 (51.6) | 101 (50.8) | 49 (48.5) | 59 (53.2) | 55 (47.0) | 114 (50.0) | 58 (53.7) | 39 (49.4) | 36 (46.8) | 75 (48.1) | 33 (50.0) |
| Age, Mean (SD) |  | 73.7 (6.6) | 74.1 (6.3) | 73.9 (6.5) | 73.5 (6.3) | 73.3 (6.9) | 74.0 (6.1) | 73.6 (6.5) | 72.9 (6.2) | 74.0 (6.94) | 73.7 (6.14) | 73.8 (6.53) | 73.6 (6.29) | 73.7 (7.2) | 73.6 (5.9) | 73.7 (6.6) | 72.9 (6.2) |
| Race, n (%) |  |  |  |  |  |  |  |  |  |  |  |  |  |  |  |  |  |
| Asian |  | 3 (1.9) | 4 (2.8) | 7 (2.3) | 2 (1.2) | 1 (0.9) | 2 (2.2) | 3 (1.5) | 1 (1.0) | 1 (0.9) | 3 ( 2.6) | 4 (1.8) | 1 (0.9) | 0 | 2 (2.6) | 2 (1.3) | 0 |
| Black or other African American |  | 0 | 0 | 0 | 2 (1.2) | 0 | 0 | 0 | 1 (1.0) | 0 | 0 | 0 | 1 (0.9) | 0 | 0 | 0 | 1 (1.5) |
| Other |  | 1 (0.6) | 0 | 1 (0.3) | 3 (1.8) | 0 | 0 | 0 | 2 (2.0) | 0 | 0 | 0 | 1 (0.9) | 0 | 0 | 0 | 1 (1.5) |
| White |  | 150 (97.4) | 140 (97.2) | 290 (97.3) | 157 (95.7) | 105 (99.1) | 91 (97.8) | 196 (98.5) | 97 (96.0) | 110 (99.1) | 114 (97.4) | 224 (98.2) | 105 (97.2) | 79 (100) | 75 (97.4) | 154 (98.7) | 64 (97.0) |
| Ethnicity, n (%) |  |  |  |  |  |  |  |  |  |  |  |  |  |  |  |  |  |
| Hispanic or Latino/a or of Spanish origin |  | 5 (3.2) | 2 (1.4) | 7 (2.3) | 1 (0.6) | 3 (2.8) | 0 | 3 (1.5) | 0 | 4 (3.6) | 2 (1.7) | 6 (2.6) | 0 | 3 (3.8) | 0 | 3 (1.9) | 0 |
| Not disclosed |  | 7 (4.5) | 6 (4.2) | 13 (4.4) | 8 (4.9) | 5 (4.7) | 6 (6.5) | 11 (5.5) | 6 (5.9) | 4 (3.6) | 5 (4.3) | 9 (3.9) | 5 (4.6) | 4 (5.1) | 5 (6.5) | 9 (5.8) | 3 (4.5) |
| Not Hispanic or Latino/a or of Spanish origin |  | 142 (92.2) | 136 (94.4) | 278 (93.3) | 155 (94.5) | 98 (92.5) | 87 (93.5) | 185 (93.0) | 95 (94.1) | 103 (92.8) | 110 (94.0) | 213 (93.4) | 103 (95.4) | 72 (91.1) | 72 (93.5) | 144 (92.3) | 63 (95.5) |
| APOE ε4 genotype, n (%) |  |  |  |  |  |  |  |  |  |  |  |  |  |  |  |  |  |
| Noncarrier |  | 47 (30.5) | 47 (32.6) | 94 (31.5) | 46 (28.0) | 30 (28.3) | 32 (34.4) | 62 (31.2) | 24 (23.8) | 37 (33.3) | 37 (31.6) | 74 (32.5) | 34 (31.5) | 24 (30.4) | 26 (33.8) | 50 (32.1) | 19 (28.8) |
| Carrier |  | 99 (64.3) | 89 (61.8) | 188 (63.1) | 106 (64.6) | 70 (66.0) | 57 (61.3) | 127 (63.8) | 73 (72.3) | 70 (63.1) | 75 (64.1) | 145 (63.6) | 71 (65.7) | 51 (64.6) | 47 (61.0) | 98 (62.8) | 45 (68.2) |
| Heterozygotes |  | 69 (44.8) | 65 (45.1) | 134 (45.0) | 76 (46.3) | 47 (44.3) | 41 (44.1) | 88 (44.2) | 52 (51.5) | 47 (42.3) | 56 (47.9) | 103 (45.2) | 45 (41.7) | 34 (43.0) | 35 (45.5) | 69 (44.2) | 30 (45.5) |
| Homozygotes |  | 30 (19.5) | 24 (16.7) | 54 (18.1) | 30 (18.3) | 23 (21.7) | 16 (17.2) | 39 (19.6) | 21 (20.8) | 23 (20.7) | 19 (16.2) | 42 (18.4) | 26 (24.1) | 17 (21.5) | 12 (15.6) | 29 (18.6) | 15 (22.7) |
| Missing |  | 8 (5.2) | 8 (5.6) | 16 (4.0) | 12 (7.3) | 6 (5.7) | 5 (5.4) | 11 (5.5) | 8 (7.9) | 4 (3.6) | 5 (4.3) | 9 (3.9) | 3 (2.8) | 4 (5.1) | 5 (6.5) | 9 (5.8) | 4 (6.1) |
| Baseline Clinical Scores, Mean (SD) |  |  |  |  |  |  |  |  |  |  |  |  |  |  |  |  |  |
| ADAS-Cog13 |  | 28.4 (8.4) | 28.9 (9.1) | 28.5 (8.5) | 30.4 (8.4) | 28.6 (8.51) | 29.3 (9.68) | 28.9 (9.06) | 29.9 (8.87) | 27.6 (8.71) | 28.8 (9.41) | 28.2 (9.07) | 30.8 (8.80) | 28.4 (9.10) | 29.3 (10.00) | 28.8 (9.54) | 30.4 (9.23) |
| ADCS-ADL |  | 66.7 (7.4) | 67 (7.9) | 66.9 (7.6) | 66.4 (7.1) | 65.9 (7.53) | 67.9 (7.42) | 66.8 (7.53) | 67.1 (6.47) | 66.3 (7.54) | 67.1 (8.01) | 66.7 (7.78) | 65.9 (7.00) | 65.3 (7.68) | 67.8 (7.71) | 66.5 (7.77) | 66.8 (6.01) |
| CDR-SB |  | 3.8 (1.6) | 3.8 (1.8) | 3.8 (1.7) | 4.1 (1.8) | 3.96 (1.708) | 3.66 (1.819) | 3.82 (1.763) | 3.95 (1.727) | 3.78 (1.755) | 3.73 (1.770) | 3.75 (1.759) | 4.22 (1.786) | 4.02 (1.853) | 3.66 (1.903) | 3.84 (1.880) | 4.02 (1.699) |
| MMSE |  | 23.6 (3.1) | 23.6 (2.8) | 23.6 (2.9) | 23.0 (2.7) | 23.6 (3.17) | 23.6 (2.84) | 23.6 (3.02) | 23.2 (2.79) | 23.7 (3.18) | 23.4 (2.85) | 23.6 (3.01) | 22.9 (2.49) | 23.5 (3.21) | 23.4 (2.96) | 23.5 (3.08) | 23.0 (2.55) |
| Baseline CDR-Global scores, n (%) |  |  |  |  |  |  |  |  |  |  |  |  |  |  |  |  |  |
| 0 |  | 0 | 1 (0.7) | 1 (0.3) | 0 | 0 | 0 | 0 | 0 | 0 | 1 ( 0.9) | 1 ( 0.4) | 0 | 0 | 0 | 0 | 0 |
| 0.5 |  | 98 (63.6) | 96 (66.7) | 194 (65.1) | 94 (57.3) | 61 (57.5) | 63 (67.7) | 124 (62.3) | 61 (60.4) | 68 (61.3) | 78 (66.7) | 146 (64.0) | 62 (57.4) | 43 (54.4) | 51 (66.2) | 94 (60.3) | 39 (59.1) |

The non-mutant populations consistently demonstrated stronger cognitive responses measured on the ADAS-Cog13 than the ITT whole group (Figures 1a-g), with ABCLEAR1 population mean difference vs. placebo of −2.317, P=0.015, representing a 49.8% reduction in clinical decline at 48 weeks [10]. For ABCLEAR2 population, mean difference vs. placebo was −3.901, P < 0.0001, representing a 59.8% reduction in clinical decline at 48 weeks. For ABCLEAR3 population, mean difference vs. placebo was −4.179, P=0.0005, representing a 75.9% reduction in clinical decline at 48 weeks. The strongest outcomes were in the ABCLEAR3 30 mg dose group (Figure 2a) with an 84.7% reduction in decline at 48 weeks, with mean difference vs. placebo of −4.739, P=0.0004 for the primary endpoint ADAS-Cog13.

**Figure 1a.**
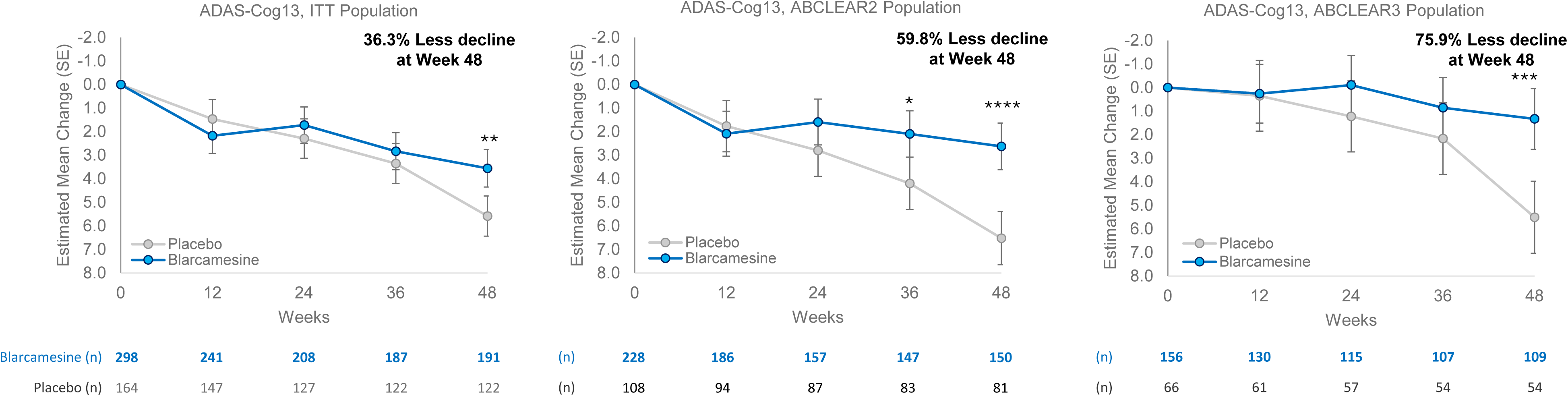
Clinical efficacy endpoint ADAS-Cog13 estimated mean change from baseline, blarcamesine versus placebo; ITT, ABCLEAR2, ABCLEAR3 populations. Clinical efficacy endpoints were analyzed using mixed model for repeated measures (MMRM) estimates for the least-squares mean change from baseline at 12, 24, 36, and 48 weeks, with error bars representing standard error (SE). The number of trial participants with analyzed results at each visit is noted beneath the x axis. Asterisks indicate statistically significant differences, where *: p < 0.05; **: p < 0.01; ***: p < 0.001; ****: p < 0.0001. “Less decline” is calculated as the percentage difference in Estimated Mean Change between placebo and active treatment groups at 48 weeks/end of study.

**Figure 1b.**
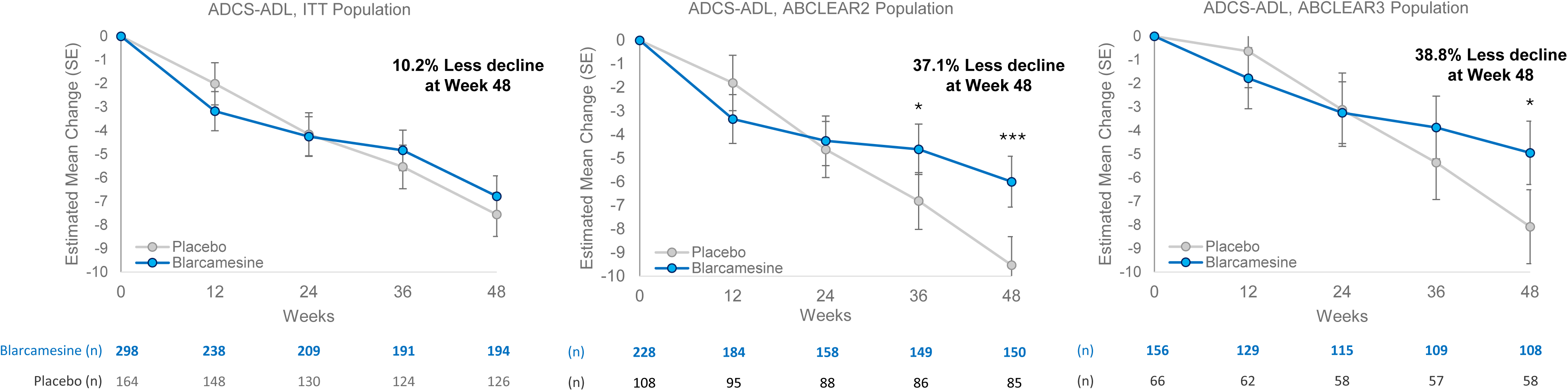
Clinical efficacy endpoint ADCS-ADL estimated mean change from baseline, blarcamesine versus placebo; ITT, ABCLEAR2, ABCLEAR3 populations. Clinical efficacy endpoints were analyzed using mixed model for repeated measures (MMRM) estimates for the least-squares mean change from baseline at 12, 24, 36, and 48 weeks, with error bars representing standard error (SE). The number of trial participants with analyzed results at each visit is noted beneath the x axis. Asterisks indicate statistically significant differences, where *: p < 0.05; **: p < 0.01; ***: p < 0.001; ****: p < 0.0001. “Less decline” is calculated as the percentage difference in Estimated Mean Change between placebo and active treatment groups at 48 weeks/end of study.

**Figure 1c.**
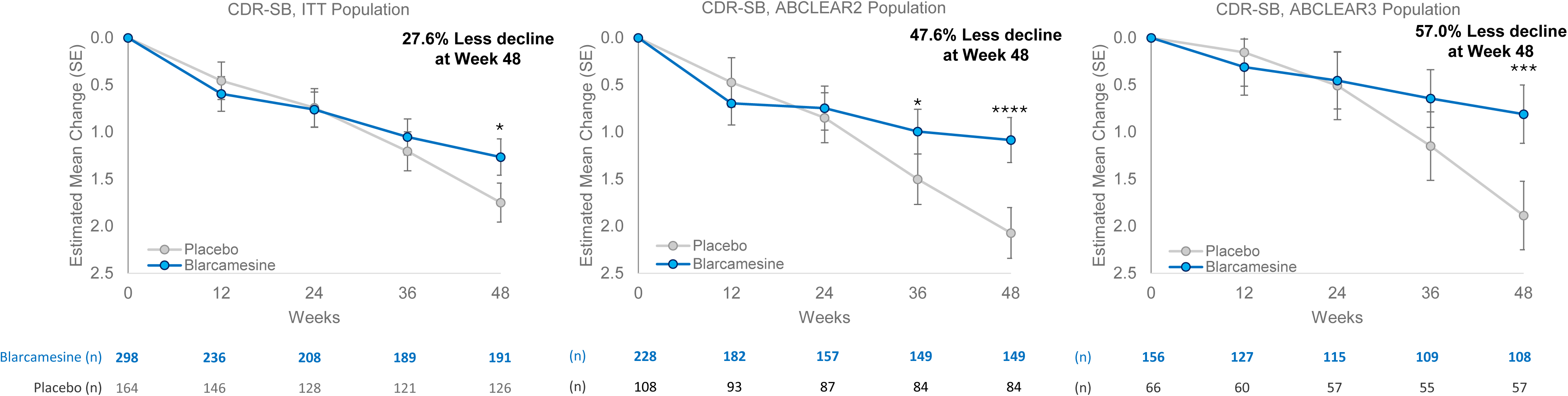
Clinical efficacy endpoint CDR-SB estimated mean change from baseline, blarcamesine versus placebo; ITT, ABCLEAR2, ABCLEAR3 populations. Clinical efficacy endpoints were analyzed using mixed model for repeated measures (MMRM) estimates for the least-squares mean change from baseline at 12, 24, 36, and 48 weeks, with error bars representing standard error (SE). The number of trial participants with analyzed results at each visit is noted beneath the x axis. Asterisks indicate statistically significant differences, where *: p < 0.05; **: p < 0.01; ***: p < 0.001; ****: p < 0.0001. “Less decline” is calculated as the percentage difference in Estimated Mean Change between placebo and active treatment groups at 48 weeks/end of study.

**Figure 1d.**
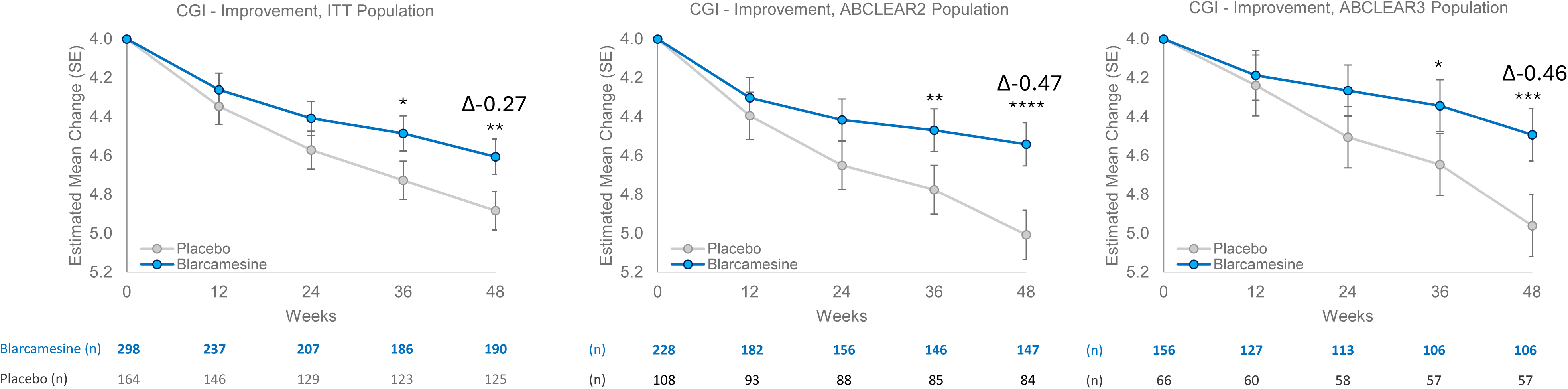
Clinical efficacy endpoint CGI-Improvement estimated mean change from baseline, blarcamesine versus placebo; ITT, ABCLEAR2, ABCLEAR3 populations. Clinical efficacy endpoints were analyzed using mixed model for repeated measures (MMRM) estimates for the least-squares mean change from baseline at 12, 24, 36, and 48 weeks, with error bars representing standard error (SE). The number of trial participants with analyzed results at each visit is noted beneath the x axis. CGI-I baseline is represented as a score of 4, which represents “no change” in clinical improvement. Asterisks indicate statistically significant differences, where *: p < 0.05; **: p < 0.01; ***: p < 0.001; ****: p < 0.0001.

**Figure 1e.**
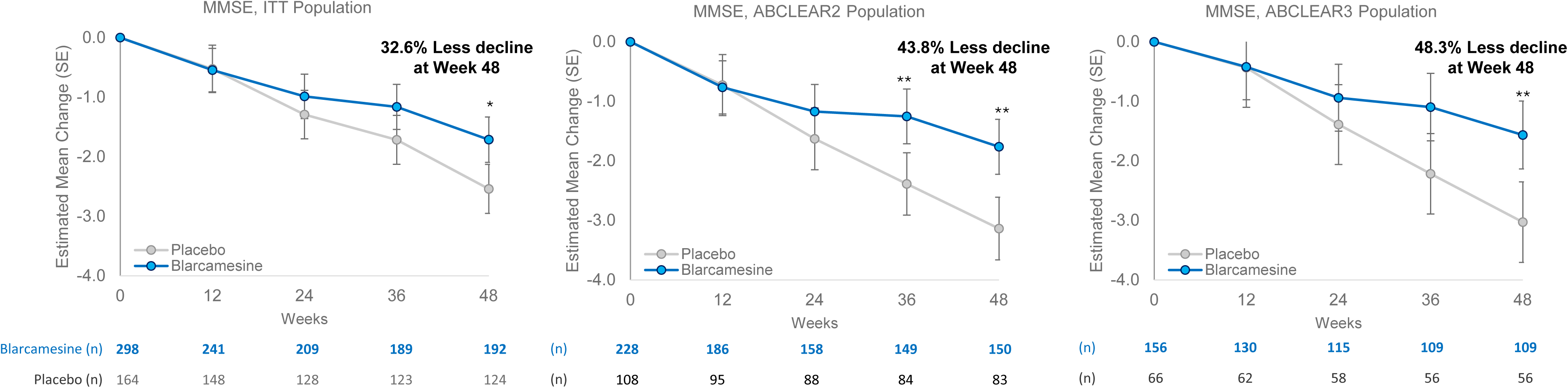
Clinical efficacy endpoint MMSE estimated mean change from baseline, blarcamesine versus placebo; ITT, ABCLEAR2, ABCLEAR3 populations. Clinical efficacy endpoints were analyzed using mixed model for repeated measures (MMRM) estimates for the least-squares mean change from baseline at 12, 24, 36, and 48 weeks, with error bars representing standard error (SE). The number of trial participants with analyzed results at each visit is noted beneath the x axis. Asterisks indicate statistically significant differences, where *: p < 0.05; **: p < 0.01; ***: p < 0.001; ****: p < 0.0001. “Less decline” is calculated as the percentage difference in Estimated Mean Change between placebo and active treatment groups at 48 weeks/end of study.

**Figure 1f.**
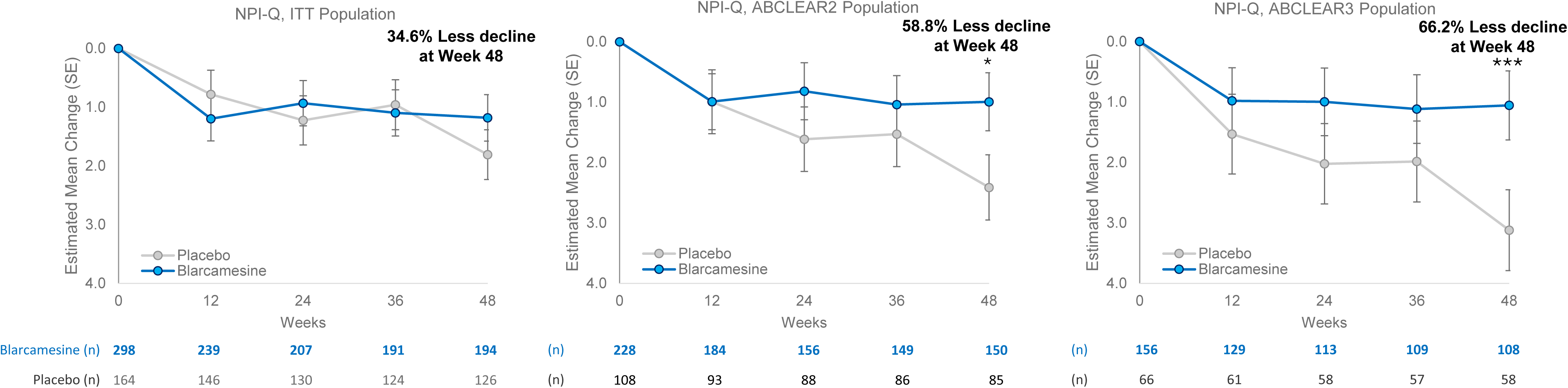
Clinical efficacy endpoint NPI-Q estimated mean change from baseline, blarcamesine versus placebo; ITT, ABCLEAR2, ABCLEAR3 populations. Clinical efficacy endpoints were analyzed using mixed model for repeated measures (MMRM) estimates for the least-squares mean change from baseline at 12, 24, 36, and 48 weeks, with error bars representing standard error (SE). The number of trial participants with analyzed results at each visit is noted beneath the x axis. Asterisks indicate statistically significant differences, where *: p < 0.05; **: p < 0.01; ***: p < 0.001; ****: p < 0.0001. “Less decline” is calculated as the percentage difference in Estimated Mean Change between placebo and active treatment groups at 48 weeks/end of study.

**Figure 1g.**
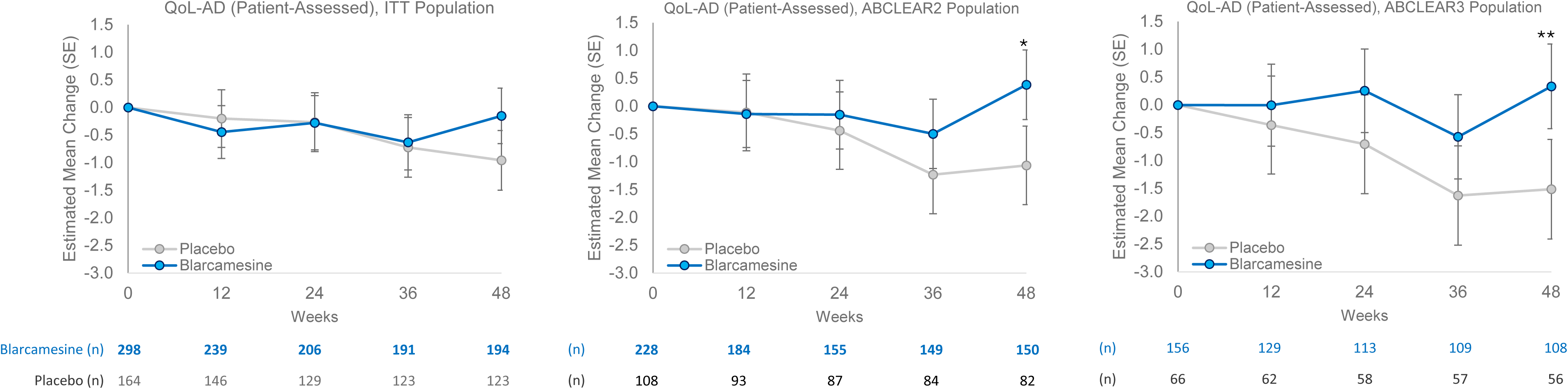
Clinical efficacy endpoint Qol-AD (Patient-Assessed) estimated mean change from baseline, blarcamesine versus placebo; ITT, ABCLEAR2, ABCLEAR3 populations. Clinical efficacy endpoints were analyzed using mixed model for repeated measures (MMRM) estimates for the least-squares mean change from baseline at 12, 24, 36, and 48 weeks, with error bars representing standard error (SE). The number of trial participants with analyzed results at each visit is noted beneath the x axis. Asterisks indicate statistically significant differences, where *: p < 0.05; **: p < 0.01; ***: p < 0.001; ****: p < 0.0001. “Less decline” is calculated as the percentage difference in Estimated Mean Change between placebo and active treatment groups at 48 weeks/end of study.

The ADCS-ADL co-primary endpoint (Figure 1b) showed marked improvement in the ABCLEAR2 group (37.1% less decline at 48 weeks) and ABCLEAR3 group (38.8% at 48 weeks). The ABCLEAR3 30 mg group had the strongest response (Figure 2a), with a 53.2% (mean difference vs. placebo of +4.245, P=0.0024) reduction in decline at 48 weeks.

**Figure 2a.**
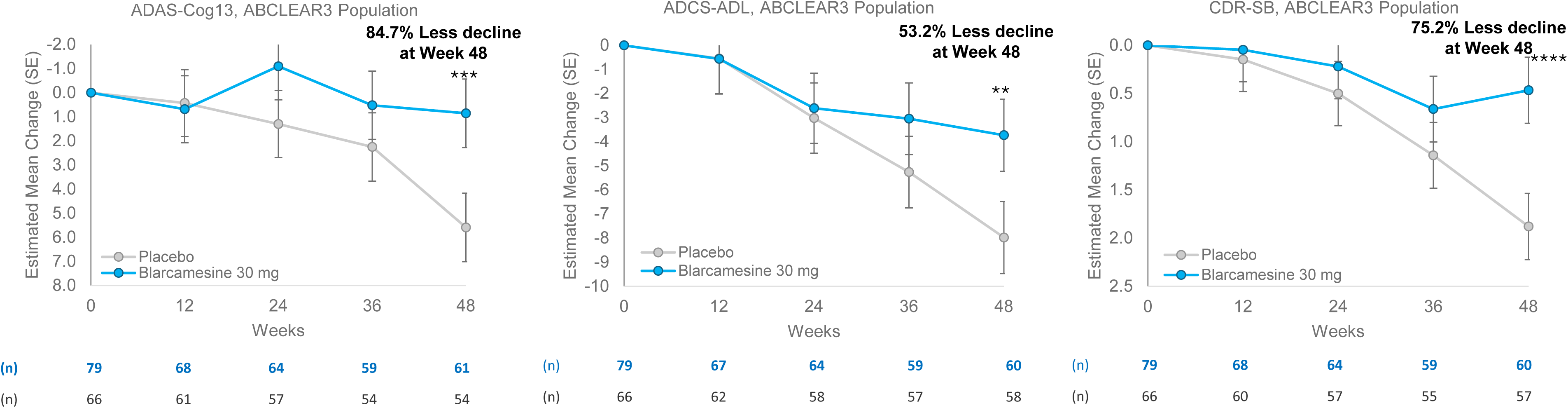
Clinical efficacy endpoints estimated mean change from baseline, blarcamesine 30 mg group versus placebo; ABCLEAR3 population. Numbers of observations below the plots represent all participants at baseline, and all participants with non-missing questionnaire and covariate data at each time point post-baseline. Values represent the least-squares mean at each analysis visit, with error bars representing standard error (SE). *: p < 0.05; **: p < 0.01; ***: p < 0.001; ****: p < 0.0001. “Less decline” is calculated as the percentage difference in Estimated Mean Change between placebo and active treatment groups at 48 weeks/end of study.

**Figure 2b.**
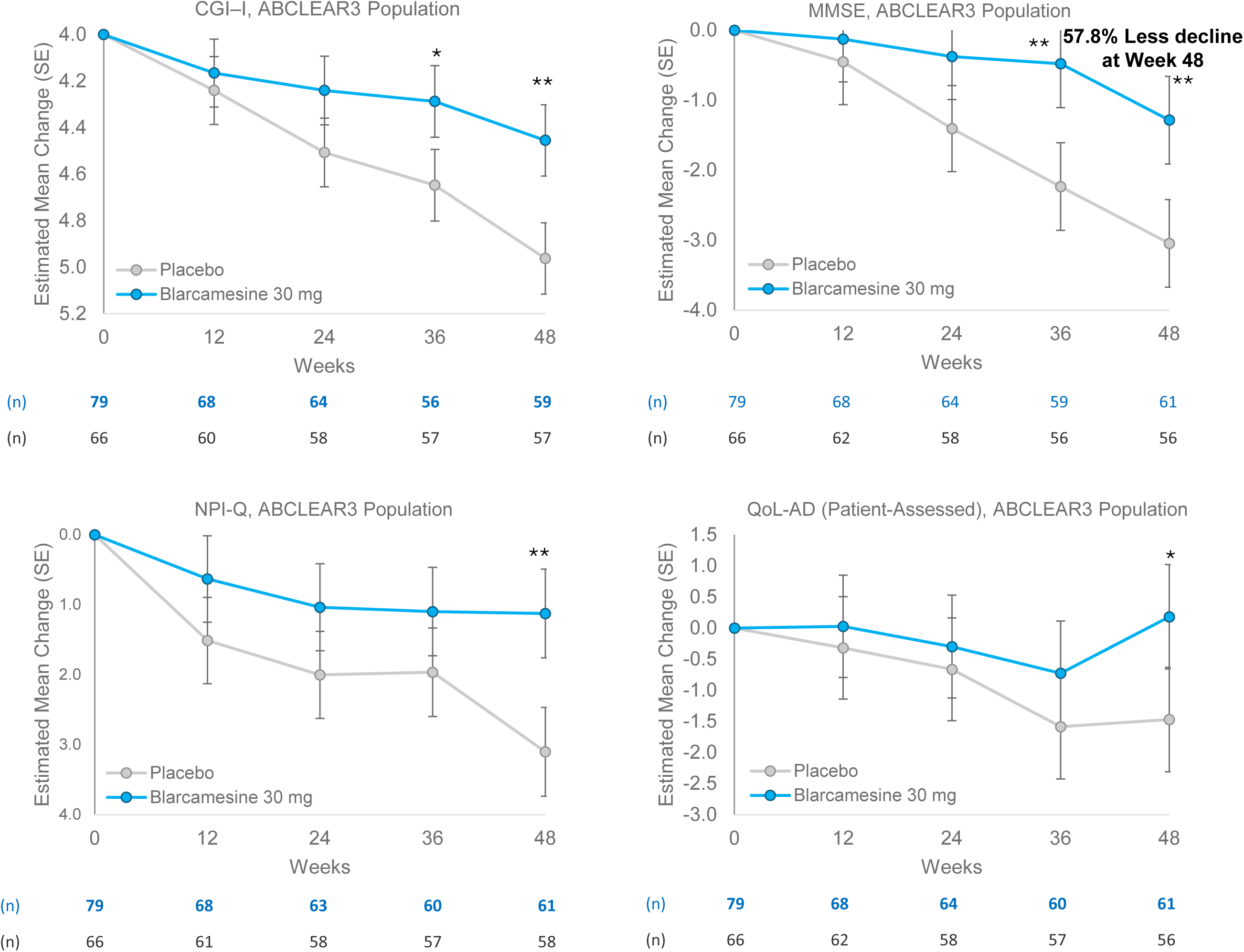
Clinical efficacy endpoints estimated mean change from baseline, blarcamesine 30 mg group versus placebo; ABCLEAR3 population. Numbers of observations below the plots represent all participants at baseline, and all participants with non-missing questionnaire and covariate data at each time point post-baseline. Values represent the least-squares mean at each analysis visit, with error bars representing standard error (SE). CGI-I baseline is represented as a score of 4, which represents “no change” in clinical improvement. *: p < 0.05; **: p < 0.01; ***: p < 0.001; ****: p < 0.0001. “Less decline” is calculated as the percentage difference in Estimated Mean Change between placebo and active treatment groups at 48 weeks/end of study.

The secondary endpoint CDR-SB (Figure 1c) had a similar pattern of progressively increasing responses comparing ITT to ABCLEAR1 population [10], ABCLEAR2 population and then ABCLEAR 3 population, from 27.6% to 57.0% reduction (mean difference vs. placebo of −1.075, P=0.0002) in decline at 48 weeks for the ABCLEAR3 population. The ABCLEAR3 30mg group had the greatest improvement of the CDR-SB (Figure 2a), mean difference vs. placebo of −1.414, P=0.0001, a 75.2% reduction in decline at 48 weeks. The respective mean differences vs. placebo of all populations are summarized in Figure 3.

**Figure 3.**
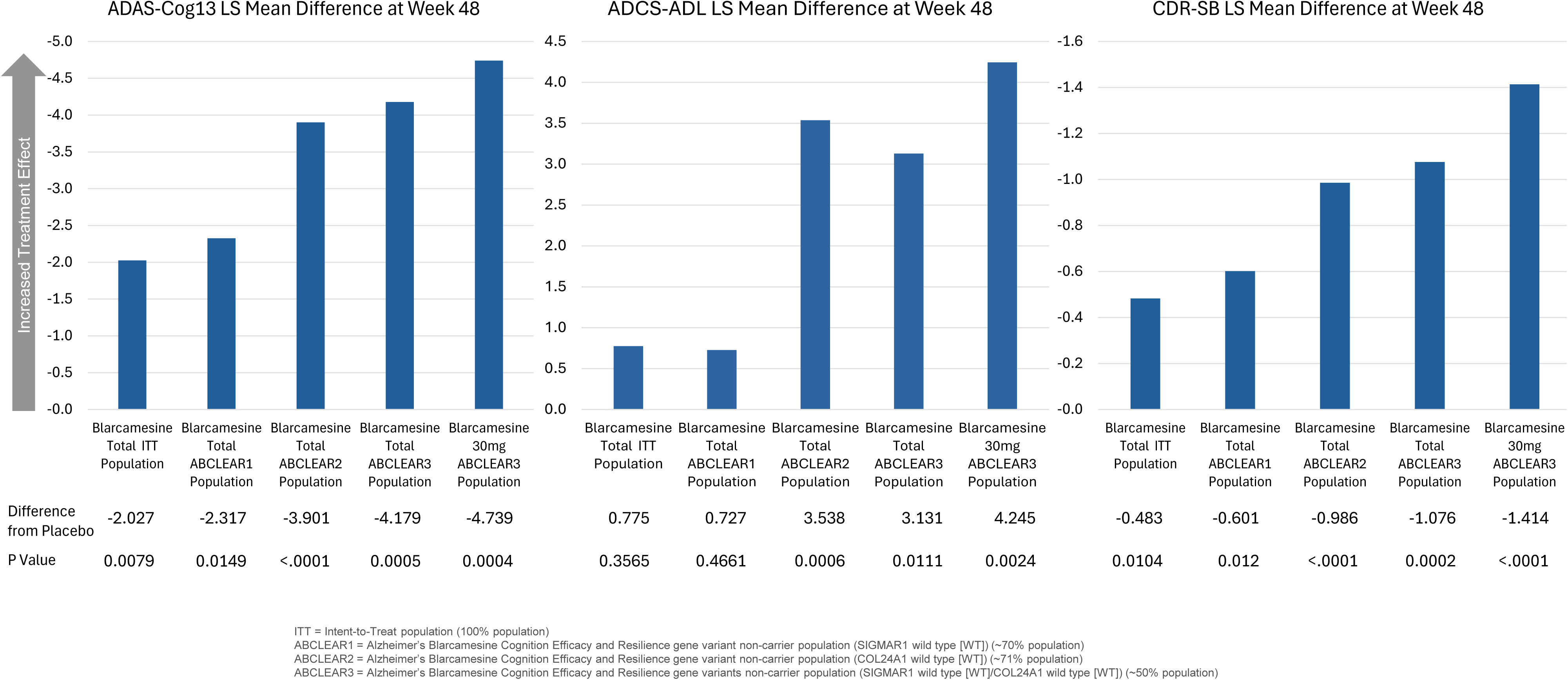
ADAS-Cog13, ADCS-ADL and CDR-SB Difference Blarcamesine vs. Placebo at Week 48

For CGI-I (Figure 1d), the ABCLEAR3 population had a strong improvement of −0.4768 (P=0.0008) at 48 weeks, with the ABCLEAR3 30 mg group having the greatest improvement (Figure 2b) with −0.508 (P=0.0014) at 48 weeks. NPI-Q results (Figure 1f) also showed sequentially deepened responses across each ABCLEAR population, with a 66.2% reduction at 48 weeks for the ABCLEAR3 population. Compared to the ITT population, MMSE (Figure 1e) decline reduction of 32.6%, the ABCLEAR3 population saw a highly significant 48.3% decline reduction at 48 weeks, with the 30 mg group achieving 57.8%, P=0.0028 at 48 weeks (Figure 2b). The patient-assessed Quality of Life (QoL-AD) scores (Figure 1g) demonstrated a significant absolute improvement in QoL-AD scores indicating a reversal of negative trajectory for AD patients from baseline to end of trial observed for blarcamesine group vs placebo; participants in the 30 mg group (Figure 2b) showed an QoL-AD LS mean change improvement from baseline score of 0.182 compared to placebo-group participants with an QoL-AD LS mean change decline from baseline score of - 1.470. The QoL-AD LS mean difference was 1.651 (95% CI 0.082, 3.221) in favor of the 30 mg group, (P=0.0392).

Effect of the apolipoprotein ε4 (APOE4) gene was also analyzed. Homozygous APOE4 ε4 influence on the treatment response in the ABCLEAR3 population was assessed for the primary endpoint ADAS-Cog13 and did not identify any imbalances of response. Within the ABCLEAR3 population, the APOE4 noncarrier cohort demonstrated an ADAS-Cog13 mean difference vs. placebo of - 6.154, P=0.0053; within the ABCLEAR3 population, for the homozygous APOE4 ε4 carriers, the ADAS-Cog13 mean treatment difference was −5.6045, P= 0.0513 (Table 2).

**Table 2.** Apolipoprotein ε4 (APOE4) Gene Analysis.

| ABCLear3<br>Population Cohort* | Week | Active | Placebo | ADAS-Cog13<br>LS Mean Diff | SE | P value | Lower 95%CI | Upper 95%CI |
| --- | --- | --- | --- | --- | --- | --- | --- | --- |
| APOE4 ε4<br>Noncarriers | 48 | 33 | 15 | -6.154 | 2.1472 | 0.0053 | -10.4246 | -1.8834 |
| APOE4 ε4<br>homozygotes | 48 | 19 | 9 | -5.6045 | 2.8196 | 0.0513 | -11.2405 | 0.03145 |

\*excludes subjects with missing ADAS-Cog13 data or covariates

Reduction in brain atrophy based on volumetric MRI data continued favorably stepwise across all populations (Figures 4a-e). For the ABCLEAR3 population, reductions in atrophy versus placebo at 48 weeks were 44.5% (P=0.0019) for the whole brain at 48 weeks (Figure 4a), and 65.2% (P=0.0013) for total grey matter at 48 weeks (Figure 4b).

**Figure 4a.**
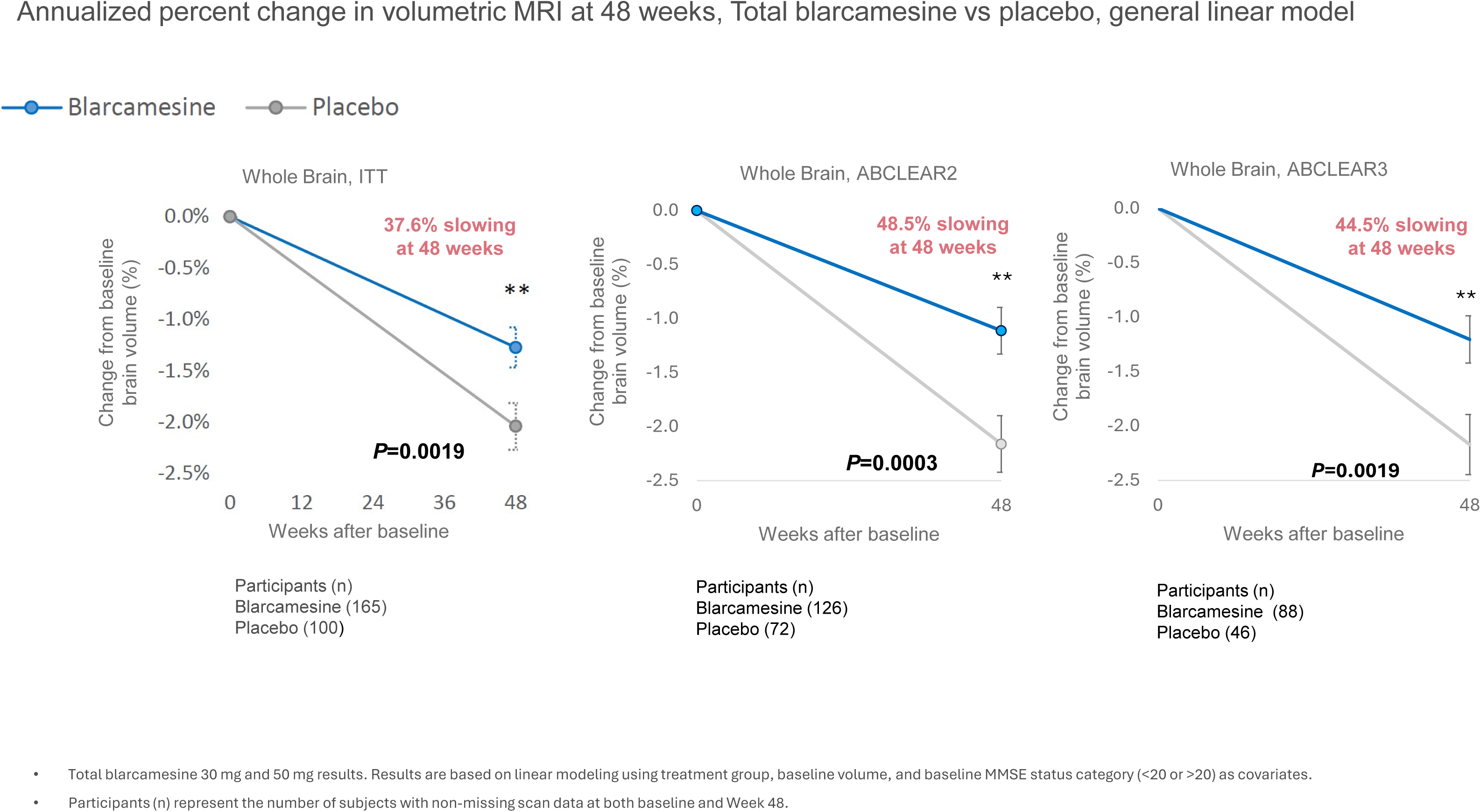
Whole Brain Atrophy in Blarcamesine-Treated Patients Compared to Placebo; ITT, ABCLEAR2, ABCLEAR3 Populations.

**Figure 4b.**
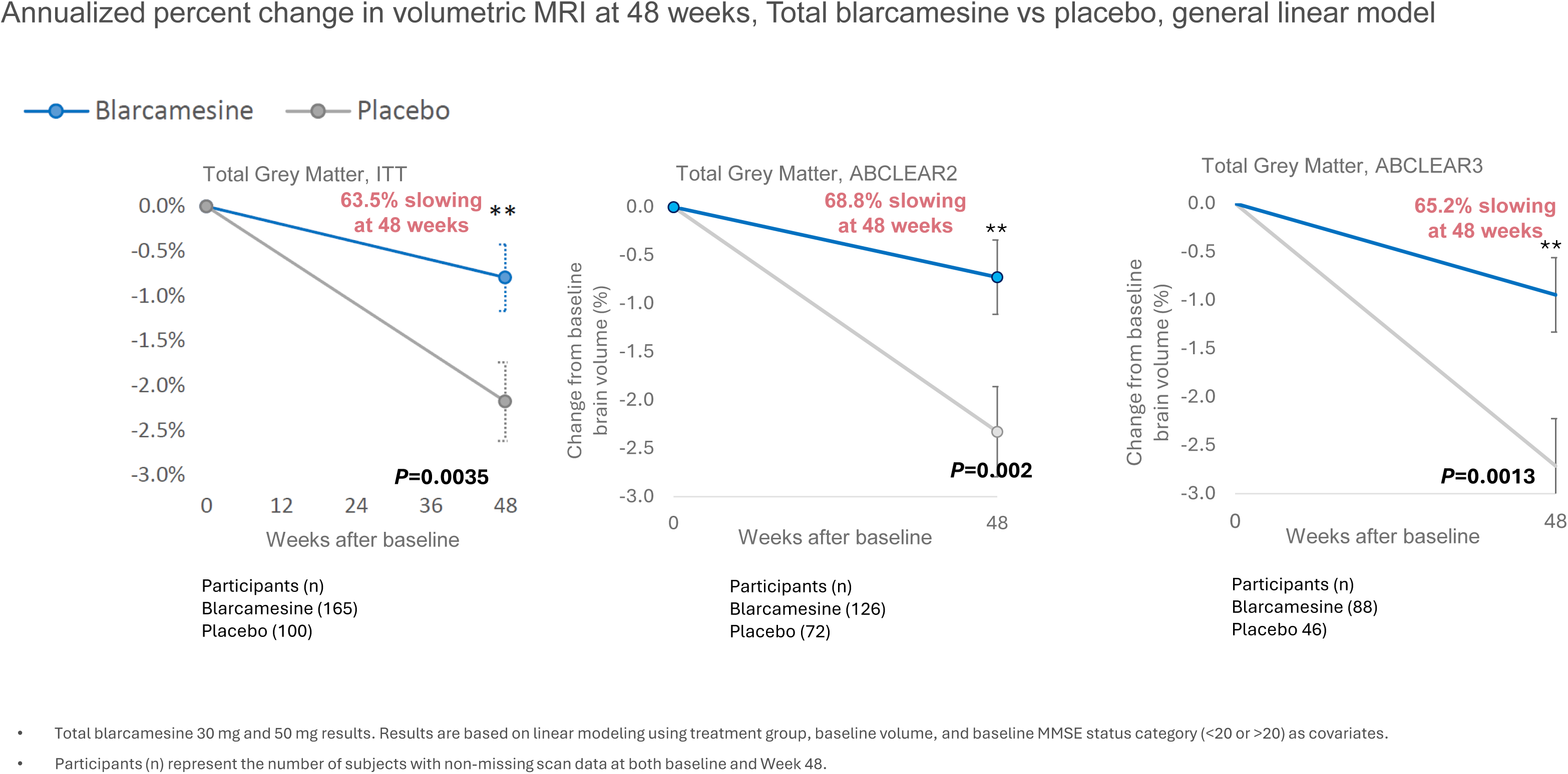
Total Grey Matter Atrophy in Blarcamesine-Treated Patients Compared to Placebo; ITT, ABCLEAR2, ABCLEAR3 Populations.

**Figure 4c.**
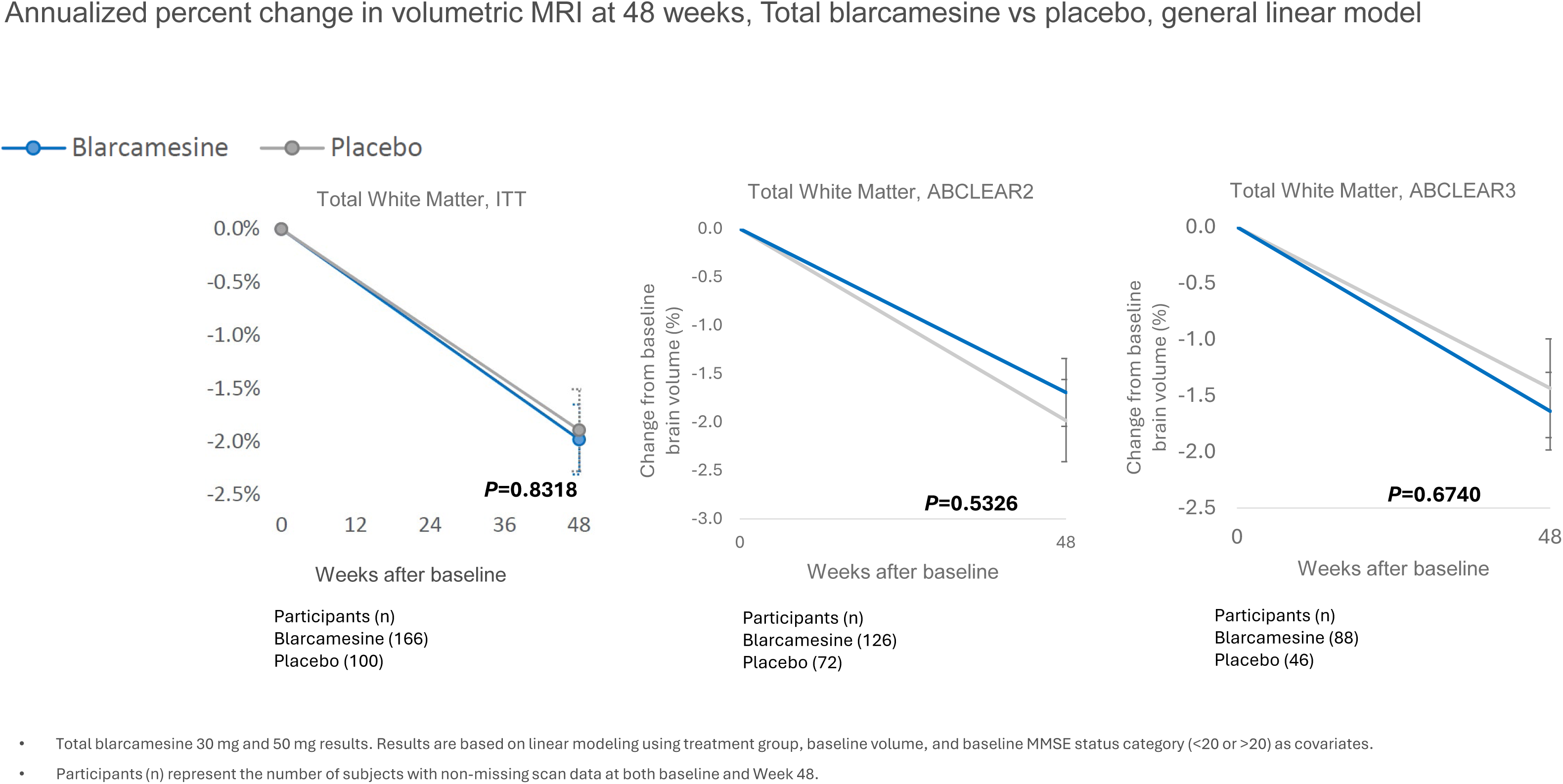
Total White Matter Brain Atrophy in Blarcamesine-Treated Patients Compared to Placebo; ITT, ABCLEAR2, ABCLEAR3 Populations.

**Figure 4d.**
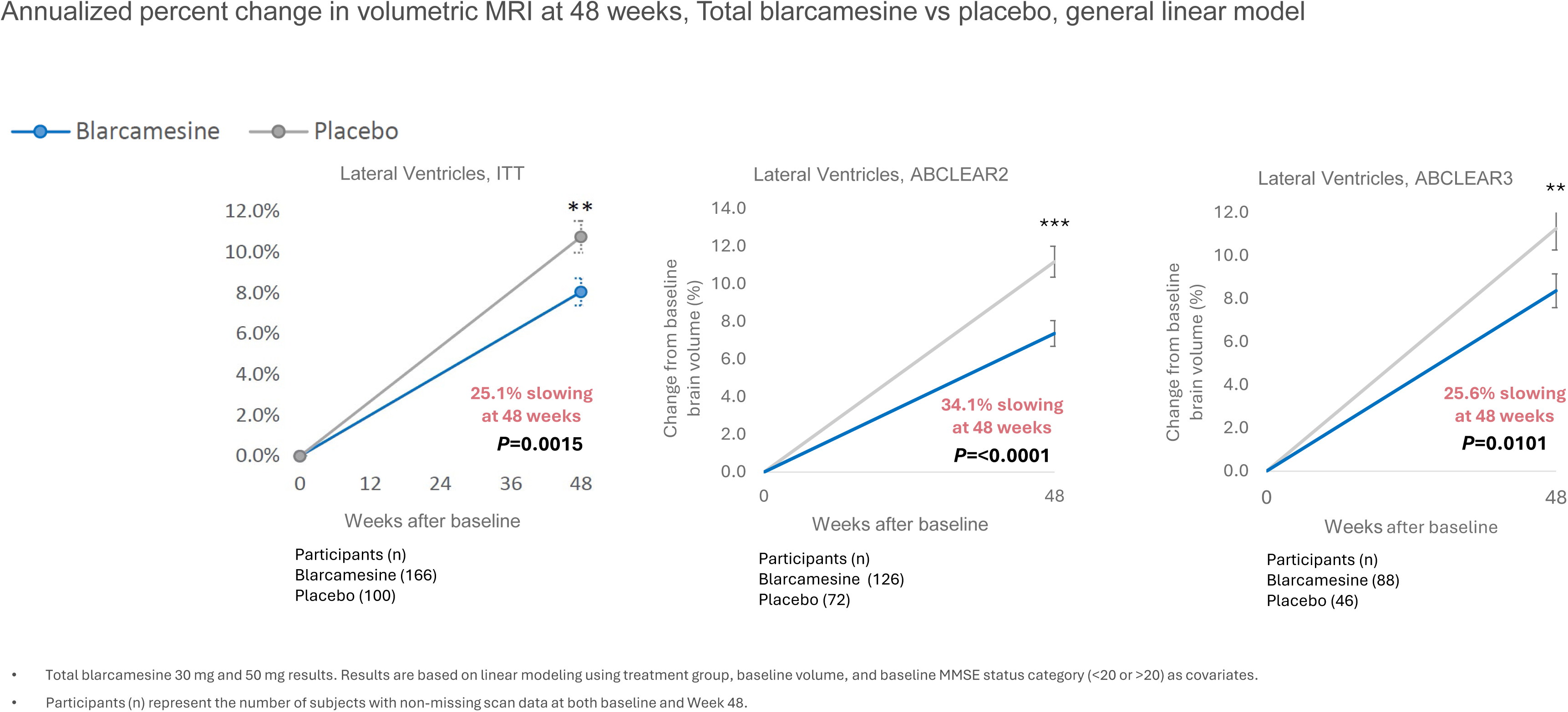
Lateral Ventricle Volume in Blarcamesine-Treated Patients Compared to Placebo; ITT, ABCLEAR2, ABCLEAR3 Populations.

**Figure 4e.**
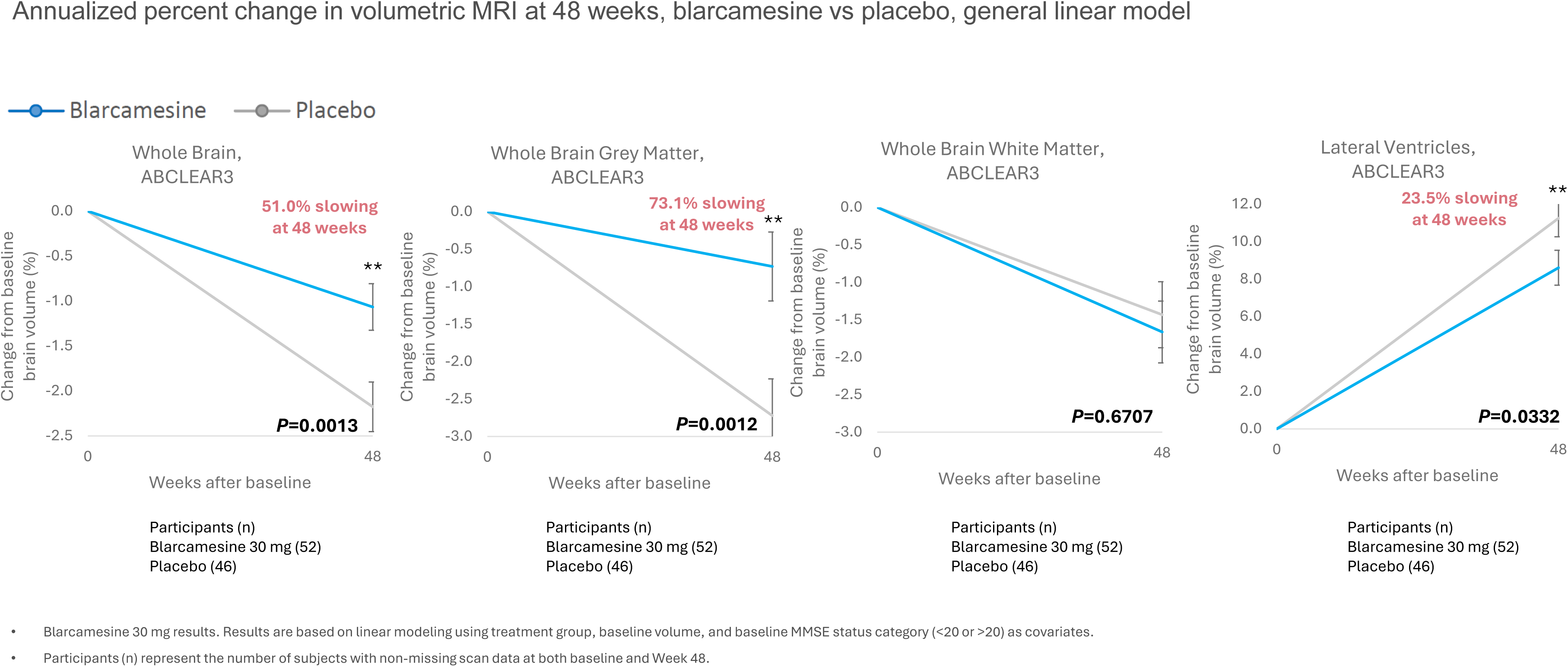
Reduced Brain Atrophy in Blarcamesine-Treated (30mg Dose Group) Patients Compared to Placebo; ABCLEAR3 Population.

For the ABCLEAR3 population, the tipping point analysis (Table 3) was performed under the missing not at random (MNAR) assumption. For the ABCLEAR3 population the tipping point shift increased to 9.74 from 1.88 (ITT population). Patients with missing data at the end of Week 48 in the active arm will have to worsen by more than 10 points in ADAS-Cog13 to invalidate the primary analysis result, which is not plausible.

**Table 3.**
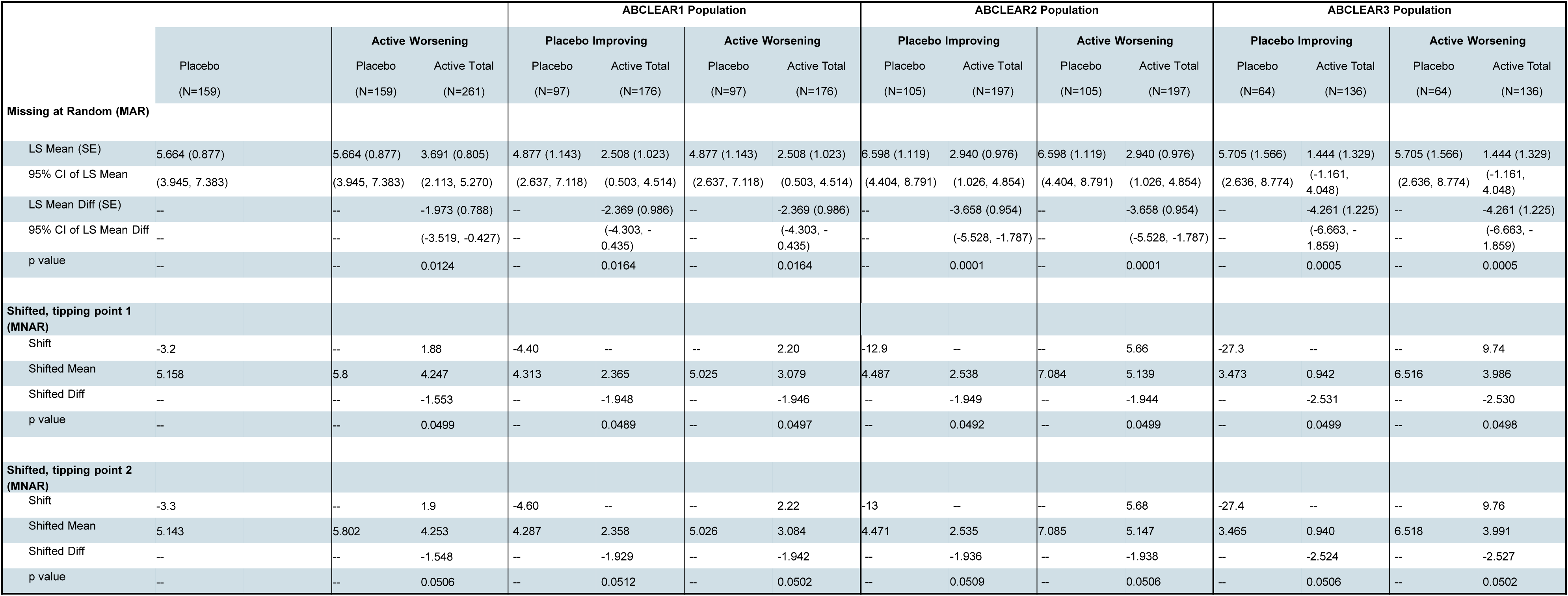
Sensitivity Analysis – Tipping Point, ADAS-Cog13.

The 30 mg group safety data indicate a favorable overall safety profile, with TEAEs trending successively downward in the ABCLEAR2 and ABCLEAR3 populations (Table 4a). Participants in the ABCLEAR3 30 mg dose cohort had 10 (12.7%) serious TEAEs versus 6 (9.1%) serious TEAEs in the placebo group. There were no deaths in the blarcamesine group and 1 in the placebo group (Table 4b).

**Table 4a.** Adverse Events – Full Safety Population and ABCLEAR Populations.

|  | Full Safety Population |  | ABCLEAR2 Population |  | ABCLEAR3 Population |  |
| --- | --- | --- | --- | --- | --- | --- |
|  | Blarcamesine Total | Placebo | Blarcamesine Total | Placebo | Blarcamesine Total | Placebo |
| **Adverse Events Summary** |  |  |  |  |  |  |
| **Participants, n** | 335 | 168 | 228 | 108 | 156 | 66 |
| Death, n (%) | 1 (0.3) | 1 (0.6) | 1 (0.4) | 1 (0.9) | 1 (0.6) | 1 (1.5) |
| Death considered related to treatment | 0 | 0 | 0 | 0 | 0 | 0 |
| Participants with ≥1 Serious TEAEs, n (%) | 56 (16.7) | 17 (10.1) | 36 (15.8) | 11 (10.2) | 28 (17.9) | 6 (9.1) |
| **TEAE, n (%)** | 324 (96.7) | 129 (76.8) | 220 (96.5) | 87 (80.6) | 151 (96.8) | 53 (80.3) |
| TEAE leading to Treatment and Study Discontinuation, n (%) | 103 (30.7) | 12 (7.1) | 57 (25.0) | 7 (6.5) | 36 (23.1) | 3 (4.5) |
| **Treatment Titration AE ≥5%, n (%)** | 335 | 168 | 228 | 108 | 156 | 66 |
| Dizziness | 120 (35.8) | 10 (6.0) | 79 (34.6) | 6 (5.6) | 54 (34.6) | 4 (6.1) |
| Confusional state | 48 (14.3) | 1 (0.6) | 36 (15.8) | 1 (0.9) | 26 (16.7) | 0 |
| Balance disorder | 25 (7.5) | 1 (0.6) | 17 (7.5) | 1 (0.9) | 10 (6.4) | 1 (1.5) |
| Fatigue | 19 (5.7) | 0 | 12 (5.3) | 0 | 6 (3.8) | 0 |
| Lethargy | 18 (5.4) | 0 | 13 (5.7) | 2 (1.9) | 9 (5.8) | 1 (1.5) |
| Anxiety | 21 (6.3) | 8 (4.8) | 10 (4.4) | 0 | 7 (4.5) | 0 |
| Headache | 48 (14.3) | 1 (0.6) | 13 (5.7) | 1 (0.9) | 10 (6.4) | 1 (1.5) |
| Nausea | 25 (7.5) | 1 (0.6) | 15 (6.6) | 7 (6.5) | 11 (7.1) | 3 (4.5) |
| **Treatment Maintenance AE ≥5%, n (%)** | 301 | 161 | 214 | 106 | 148 | 65 |
| Dizziness | 76 (25.2) | 9 (5.6) | 61 (28.5) | 5 (4.7) | 44 (29.7) | 2 (3.1) |
| Confusional state | 40 (13.3) | 4 (2.5) | 25 (11.7) | 3 (2.8) | 18 (12.2) | 2 (3.1) |
| Fall | 21 (7.0) | 16 (9.9) | 16 (7.5) | 10 (9.4) | 11 (7.4) | 6 (9.2) |
| Depressed mood | 15 (5.0) | 3 (1.9) | 12 (5.6) | 2 (1.9) | 6 (4.1) | 1 (1.5) |
| Headache | 19 (6.3) | 6 (3.7) | 17 (7.9) | 2 (1.9) | 10 (6.8) | 2 (3.1) |
| Anxiety | 17 (5.6) | 6 (3.7) | 11 (5.1) | 5 (4.7) | 8 (5.4) | 3 (4.6) |
| Balance Disorder | 16 (5.3) | 2 (1.2) | 15 (7.0) | 2 (1.9) | 10 (6.8) | 1 (1.5) |
| Disorientation | 15 (5.0) | 3 (1.9) | 9 (4.2) | 0 | 8 (5.4) | 0 |

ABCLEAR1 = Alzheimer's Blarcamesine Cognition Efficacy and Resilience gene variant non-carrier population (SIGMAR1 wild type [WT]) – relevant data provided in [10]
ABCLEAR2 = Alzheimer's Blarcamesine Cognition Efficacy and Resilience gene variant non-carrier population (COL24A1 wild type [WT])
ABCLEAR3 = Alzheimer's Blarcamesine Cognition Efficacy and Resilience gene variants non-carrier population (SIGMAR1 wild type [WT]/COL24A1 wild type [WT])

**Table 4b.**
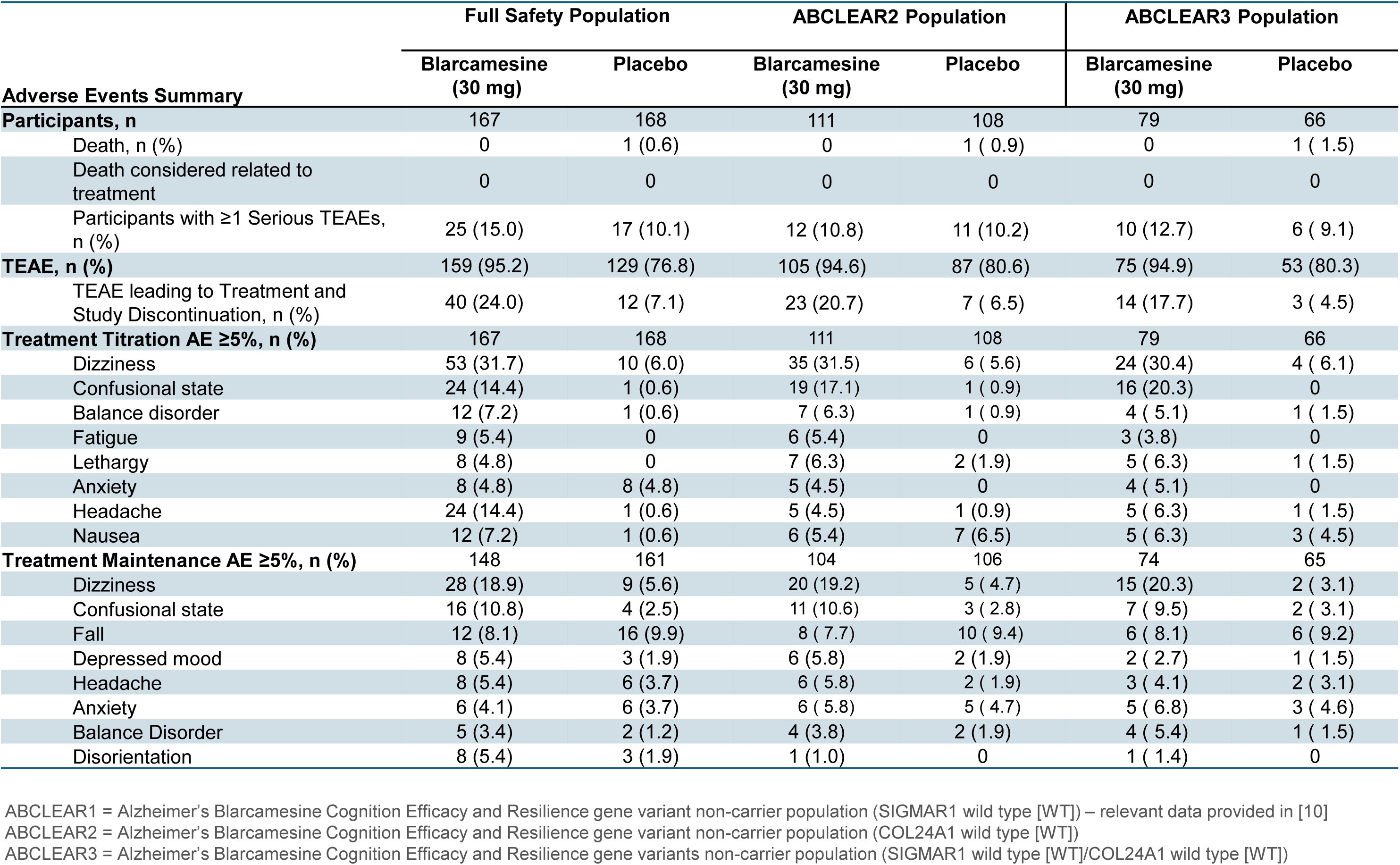
Adverse Events – Full Safety Population and ABCLEAR Populations – 30 mg Blarcamesine.

Trial participants were offered the option to continue blarcamesine for up to Week 192 in an OLE study, generating evidence of a longer duration of potential efficacy and safety, which will be reported later. However, in summary, continued blarcamesine treatment—without interruption—is encouraged for continued favorable clinical outcomes. OLE results show the importance of continued long-term blarcamesine treatment and the importance of early intervention that may indicate a disease-modifying effect. [48, 49]

## DISCUSSION

Here we provide for the first time evidence of Precision Medicine neurology treatment responses with a once-daily oral therapeutic intervention in early AD, with a differentiated upstream and constitutional mechanism of action utilizing prespecified and GWAS identified genetic variants SIGMAR1 (ABCLEAR1), COL24A1 (ABCLEAR2) and SIGMAR1/COL24A1 (ABCLEAR3) populations in a placebo-controlled clinical trial of 48 weeks. The resulting robust clinical responses indicate strong potential to change the clinical disease course. In addition, to previously confirmed prespecified Precision Medicine patient benefit in the ABCLEAR1 population compared to the ITT population [10], blarcamesine demonstrated significant improvement of all prespecified clinical endpoints, including ADAS-Cog13, ADCS-ADL, CDR-SB, MMSE, CGI-I, NPI-Q and QoL-AD across the genetically defined ABCLEAR populations. Furthermore, blarcamesine was associated with a significant reduction in MRI measures of brain atrophy, indicating an impact on underlying neurodegeneration. The safety profile remained favorable and consistent across all ABCLEAR populations, with ABCLEAR3 population demonstrating further improvement of the already adequate safety profile.

All clinical endpoints demonstrated improvement in the blarcamesine treated group as well as the respective 30 mg and 50 mg blarcamesine groups at 48 weeks. Generally, the 30 mg dose group showed better efficacy and tolerability than the 50 mg dose group. This is likely related to the higher power retained in the 30 mg blarcamesine group coupled will further improved safety profile. Overall, this study demonstrated the 30 mg blarcamesine dose as having the most balanced benefit-to-risk ratio. In aggregate, the clinical endpoints demonstrate efficacy based on current regulatory standards for early AD, and the magnitudes of the clinical effects demonstrate solid efficacy which appears to compare favorably to recently approved therapies for early AD. Importantly, blarcamesine’s favorable safety profile and ease of administration position it as a practical alternative to infusion-based monoclonal antibody therapies, which are burdened by ARIA risks and logistical complexity.

APOE4 influence on the treatment response in the ABCLEAR3 population was assessed and did not identify any imbalances of response. This fact might be advantageous in geographic regions, where patients, who are APOE4 homozygous carriers, and hence are most in need for a treatment, are precluded from receiving recently approved injectable monoclonal antibodies.

The results of the prespecified ABCLEAR1 gene population analysis reinforce the previously confirmed (from the earlier published Phase 2a AD study) [17] mechanism of action for blarcamesine in AD, beneficial clinical effect through upstream SIGMAR1 activation. Compared to the full ITT population, subjects *without* the missense SIGMAR1 (rs1800866) mutation, i.e. including the common SIGMAR1 WT carriers; n = 199/101 blarcamesine/placebo, treated with blarcamesine experienced a greater clinical benefit for both ADAS-Cog13 (slowed clinical progression by 49.8% vs. 36.3%) and CDR-SB (slowed clinical progression by 33.7% vs. 27.6%). The respective clinical responses were further markedly improved in the respective ABCLEAR2 and ABCLEAR3 populations. Individuals with the COL24A1 WT gene—present in 71.7% of the global population—show further improved response to blarcamesine treatment, enabling effective autophagy restoration via stabilization of the brain’s extracellular matrix. COL24A1 is broadly expressed in neurons, with highest levels in the hippocampus and cortex. WT carriers respond better than those with the His641Arg missense mutation. Further mechanistic details will be published separately. ADAS-Cog13 (slowed clinical progression by 59.8% vs. 36.3%) and CDR-SB (slowed clinical progression by 47.6% vs. 27.6%) for ABCLEAR2 population and ADAS-Cog13 (slowed clinical progression by 75.9% vs. 36.3%) and CDR-SB (slowed clinical progression by 57.0% vs. 27.6%) for the ABCLEAR3 population. The clinical outcomes were strongest in the ABCLEAR3 blarcamesine 30 mg cohort with ADAS-Cog13 slowed clinical progression by 84.7% vs. 36.3% and CDR-SB slowed clinical progression by 75.2% vs. 27.6%.

Blarcamesine Precision Medicine findings of the 30 mg cohort demonstrated cognitive outcomes to match barely detectable prodromal AD decline patterns across key standardized assessment scales. ADAS-Cog13: Blarcamesine showed a change from baseline of 0.853 compared to ∼1 point typical annual decline in prodromal AD aging adults. CDR-SB: Blarcamesine demonstrated a change from baseline of 0.465, aligning with the 0-0.5 point annual range seen in prodromal AD aging. These data are similar to referenced barely detectable prodromal AD decline, in spite of the more advanced stage of AD impairment at baseline of the blarcamesine population (MMSE, mean [SD] was 23.5 [3.21] for blarcamesine ABCLEAR3 population and 25.68 [2.2] for prodromal AD population) [50]. Oral blarcamesine treatment over 48 weeks in a Precision Medicine population, including up to ∼70% of the global population, indicate the ability to shift the cognitive decline of an early AD (comprising MCI or mild AD) patient population to a prodromal AD population. This is consistent with the 84.7% reduction in decline at 48 weeks of blarcamesine treatment vs placebo on the prespecified co-primary cognitive endpoint ADAS-Cog13 in the ABCLEAR3 population as well as significant absolute improvement in Quality of Life from baseline, indicating a reversal of the QoL trajectory for AD patients from baseline to end of trial, as observed in the placebo arm. [51]

The study had some missing data. 43 out of 462 ITT patients dropped out before reaching Week 12, the first analysis visit. Among these patients, 38 were in the blarcamesine group, and 35 dropped out due to TEAEs. The missing data in the study dropouts were primarily due to patients who did not tolerate the relatively short and steep titration schedule of this study. There is no evidence that these patients introduced a bias in favor of the blarcamesine group by dropping out early. Going forward, the titration schedule can be adjusted to slower titration and lower target dose as it has been demonstrated in the OLE ATTENTION-AD trial [48], and conforms with general principles of pharmacologic dosing in elderly patients. The ATTENTION-AD trial demonstrated the manageable nature of the most frequent treatment emergent adverse event (TEAE) of dizziness observed in the preceding ANAVEX2-73-AD-004 trial, which was generally transient in duration (approx. 7-11 days) and mild or moderate in severity (Grade 1 or 2). The titration schedule was adjusted to a slightly longer titration period in the ATTENTION-AD trial, from previous 2-3 weeks to 10 weeks. A markedly lower frequency of the TEAE of dizziness in the respective maintenance phase was observed: from previously 25.2% in the ANAVEX2-73-AD-004 trial to 9.6% in the ATTENTION-AD trial, demonstrating the manageable nature of the most frequent TEAE (dizziness).

To our knowledge this is the first report of a therapeutic agent for AD that has demonstrated an attenuation in global brain volume loss measured by MRI and reduction of the expansion of the lateral ventricular volume compared to placebo in the ITT population, which was further improved in the respective ABCLEAR2 and ABCLEAR3 populations. Volumetric MRI improvements associated with blarcamesine appeared global and may be in response to restoration of cellular homeostasis [16]. The global reductions in volumetric MRI atrophy rates observed with blarcamesine are associated with a marked attenuation of clinical disease progression, suggesting that the therapeutic effects may be mediated though mitigation of ongoing neurodegeneration. In contrast, anti-amyloid beta monoclonal antibodies have been associated with amyloid-related imaging abnormalities-edema (ARIA-E), amyloid-related imaging abnormalities-hemorrhages (ARIA-H) and importantly a decrease in whole brain volume, i.e. brain atrophy (ARIA-A), compared with placebo and a mean increase in ventricular volume compared with placebo [46, 47].

Blarcamesine was relatively safe in the study population, with no trends of severe or life-threatening and with no associated neuroimaging adverse events. There were no deaths attributable to blarcamesine. The initially observed early discontinuations and adverse events might be related to the timing of the up titration of blarcamesine to the target doses coupled with administration at consistent timepoints relatively early in the morning as specified in the protocol. These events can likely be addressed by changing administration to nighttime (or generally more flexible) dosing, as has been positively observed in the compassionate use program of blarcamesine administration, coupled with once daily oral dosing without requiring reaching the higher target doses. It is noteworthy that respective once-daily oral 30 mg blarcamesine cohorts in the respective ABCLEAR1, ABCLEAR2, and ABCLEAR3 populations demonstrates further improvement of the already adequate safety profile of the ITT population.

This study has some limitations. First, there was variability in total blarcamesine doses received and/or duration of blarcamesine dosing. Second, data collection was for 48 weeks, limiting long-term understanding of blarcamesine; however, a 96/144-week OLE extension study (ATTENTION-AD) followed [48]. Third, the studied populations were primarily White (96.8%), which may limit generalizability to other populations due to a lack of racial and ethnic diversity. In order to demonstrate effectiveness in a broader population, future studies will require a more diversified patient cohort. Fourth, although no related protocol amendments were necessary, this trial was conducted during the COVID-19 pandemic.

Blarcamesine, a small molecule administered orally once daily, had already demonstrated numerically superior clinical efficacy to approved therapies while also slowing neurodegeneration in early AD patients in the ITT population. Blarcamesine has a demonstrated safety profile and does not require routine MRI monitoring, and given its differentiated mechanism of action, within a heterogeneous AD population, clinical utility of blarcamesine could be enhanced via a Precision Medicine approach of treating those with target-relevant genetic profiles, excluding missense gene populations for still up to ∼70% of the early AD population. Given the robust clinical cognitive and functional response and added Quality of Life for the AD participants in the study, paired with blarcamesine’s enhanced safety profile, this effective targeting could alleviate a significant medical and economic burden.

## Data Availability

All data produced in the present study are available upon reasonable request to the authors

## Mixed competing interests

All authors have completed the ICMJE uniform disclosure form at www.icmje.org/coi_disclosure.pdf and declare: no support from any organization for the submitted work; MNS has received consulting fees from Anavex Life Sciences, Eisai, Signant, Neurotherapia, AbbVie, FujiRebio, Cognito, Novo Nordisk and Johnson & Johnson; honoraria from Lilly; stock options from Alzheon, uMETHOD and Lighthouse Pharmaceuticals; participated in a monitoring or advisory board for Bristol Myers Squibb; has a leadership role in CervoMed; SM has received medical monitoring fees for Anavex’s Rett syndrome studies; medical monitoring fees from Lilly, Johnson & Johnson and Eisai; TG received institutional support from Biogen, Eisai and Lilly; consulting fees from Acumen, Advantage Therapeutics, Alector, Anavex Life Sciences, Biogen, Bristol Myers Squibb, Cogthera, Eisai, Functional Neuromodulation, Grifols, Johnson & Johnson, Lilly, Neurimmune, Noselab, Novo Nordisk, Roche Diagnostics and Roche Pharma; honoraria from Anavex Life Sciences, Cogthera, Eisai, FEO, Grifols, Lilly, Pfizer, Roche Pharma, Schwabe and Synlab; TJO’s institution has received consulting fees and honoraria from Eisai, UCB Pharma, ES Therapeutics, Kinoxis Pharmaceuticals, Supernus, Autobahn, Epidarex, Jazz Therapeutics, Kinoxis Therapeutics and NaviFUS; travel support from Longboard Pharmaceuticals and Bright Minds Pharmaceuticals; participated in a monitoring or advisory board for Kinoxis Pharmaceuticals and Bright Minds Biopharmaceuticals; MW has received consulting fees from Cerecin; honoraria from Eisai, Roche and Lilly; travel support from Merck and Roche; BJB has received consulting fees from Eisai; CS has received honoraria from Lilly and Roche; participated in advisory boards for Eisai and Lilly; SK received an honorarium from Roche; RL has received consulting fees from Eisai; CK received research funding, consulting fees and honoraria from Roche Diagnostics; NP has received consulting fees from Novartis, Aribio and Lilly; honoraria from Lilly; LF has received institutional research support from Roche; consulting fees from Anavex Life Sciences, Biogen, BioVie, Bristol Myers Squibb, Charles River Associates, Lilly, Eisai, GE Healthcare, Grifols, Johnson & Johnson, Neurimmune, Noselab, Novo Nordisk, Roche, TauRx and Schwabe; honoraria from Lilly, Eisai, DerCampus, Medscape, Medfora, FOMF, Novo Nordisk, Roche and Schwabe; participated in a monitoring or advisory board for Neuroscios, ReMynd, Otsuka/Avanir and Vivoryon; PT received honoraria from Ipsen Pharma and Merz Therapeutics; travel support from Boston Scientific, Medtronic, Ipsen and Merz; OP has received consulting fees from Biogen, Eisai, Grifols, Noselab, Novo Nordisk, Prinnovation and Roche; honoraria from Eisai, Lilly and Roche; AHB received institutional research support from Novo Nordisk, AriBio, Alnylam, Cerevel, IGC Pharma; consulting and advisory board fees from Eisai and Lilly; honoraria from Eisai and Lilly; SHP received institutional research support from and has patent authorship with Zywie Bio; honoraria from Lilly; HC has received research site support or fees from Roche, TauRx, Lilly, Anavex Life Sciences, Alector, Biogen, Eisai and Immunocal; participated in advisory boards for Lilly, Eisai and Biogen; GRH has received institutional research support from Biogen, Roche, Cassava and Eisai; has received consulting fees from Biogen, Roche, Novo Nordisk, Eisai and Lilly; CT has received institutional research support from Johnson & Johnson, UCB, Novo Nordisk, Bristol Myers Squibb, Passage Bio, Aribio and Anavex Life Sciences; received consulting fees from Eisai, Lilly and Novo Nordisk; SC has received institutional research support from Acumen, AgeneBio, Alector, Alnylam, Alzheon, Anavex Life Sciences, Biogen, Bristol Myers Squibb, Cassava, Eisai, Eli Lilly, GAP, GSK, INmune Bio, Johnson & Johnson, Merck, Novartis, Novo Nordisk, RetiSpec, Roche, UCB, Vielight and Voyager; consulting fees from AbbVie, Biogen, Biohaven, Bristol Myers Squibb, Eisai, Kisbee Therapeutics, Lilly, Novo Nordisk, Parexel and Retispec; travel support from Biogen, Eisai, Lilly and Novo Nordisk; participated in monitoring or advisory boards for Altoida, Biogen, Cassava, Cognivue, CogState, Eisai, GSK, Lilly, Johnson & Johnson, Medscape, Novo Nordisk, Novartis, Roche and Sci Neuro. OC and EG are QYNAPSE employees with ownership interest. LV and NG are QYNAPSE employees. JE, TK, JCL, KJ and WC are employees of Anavex Life Sciences with ownership interest. DG is a consultant of Anavex Life Sciences. WL is a consultant of Anavex Life Sciences; receives intermittent consulting fees from Novartis, Google Ventures, Structure Therapeutics and Enveda; is a member of the scientific advisory boards of Axonis Therapeutics, MuriPhys and AfaSci. CM is an employee of Anavex Life Sciences with ownership interest and patent authorship. AG is a member of the scientific advisory board of Anavex Life Sciences.

## Additional Contributions

We thank all the trial participants and their families and caregivers who participated in the ANAVEX2-73-AD-004 trial as well as the site staff, raters, and site investigators; members of the data and safety monitoring board.

## SUBMISSION CATEGORY

Original Research

